# Ultra-low coverage genome-wide association study – insights into gestational age using 17,844 embryo samples with preimplantation genetic testing

**DOI:** 10.1101/2022.06.15.22276464

**Authors:** Shumin Li, Bin Yan, Thomas K.T. Li, Jianliang Lu, Yifan Gu, Yueqiu Tan, Fei Gong, Tak-Wah Lam, Pingyuan Xie, Yuexuan Wang, Ge Lin, Ruibang Luo

**Author notes:** To whom correspondence should be addressed: Ruibang Luo, Ge Lin,; Yuexuan Wang, Pingyuan Xie, Email! These authors contributed equally to this work (Shumin Li, Bin Yan, Thomas K.T. Li).

## Abstract

**Background:** Very low coverage (0.1 to 1x) whole genome sequencing (WGS) has become a promising and affordable approach to discover genomic variants of human populations for Genome-Wide Association Study (GWAS). To support genetic screening using Preimplantation Genetic Testing (PGT) in a large population, the sequencing coverage goes below 0.1x to an ultra-low level. However, its feasibility and effectiveness for GWAS remains undetermined.

**Methods:** We devised a pipeline to process ultra-low coverage WGS data and benchmarked the accuracy of genotype imputation at the combination of different coverages below 0.1x and sample sizes from 2,000 to 16,000, using 17,844 embryo PGT with approximately 0.04x average coverage and the standard Chinese sample HG005 with known genotypes. We then applied the imputed genotypes of 1,744 transferred embryos who have gestational ages and complete follow-up records to GWAS.

**Results:** The accuracy of genotype imputation under ultra-low coverage can be improved by increasing the sample size and applying a set of filters. From 1,744 born embryos, we identified 11 genomic risk loci associated with gestational ages and 166 genes mapped to these loci according to positional, expression quantitative trait locus and chromatin interaction strategies. Among these mapped genes, *CRHBP, ICAM1* and *OXTR* were more frequently reported as preterm birth related. By joint analysis of gene expression data from previous studies, we constructed interrelationships of mainly *CRHBP, ICAM1, PLAGL1, DNMT1, CNTLN, DKK1* and *EGR2* with preterm birth, infant disease and breast cancer.

**Conclusions:** This study not only demonstrates that ultra-low coverage WGS could achieve relatively high accuracy of adequate genotype imputation and is capable of GWAS, but also provides insights into uncovering genetic associations of gestational age trait existed in the fetal embryo samples from Chinese or Eastern Asian populations.

## Background

Detection and characterization of genetic variants associated with traits and diseases are fundamental to the study of human genetics. Genome-Wide Association Study (GWAS) is an approach widely used in genetic research that aims to decode the associations of specific genetic variations with particular diseases or traits in sample populations. In the past decade, GWAS has facilitated discovery of over one hundred thousand variants associated with complex traits in human (1). Whole-genome sequencing (WGS) has emerged as a dominant technology in GWAS because it enables one to generate a comprehensive view of the genomic variation landscape for not only a specific trait but also for common diseases. Thus, WGS-based approaches hold a significant advantage over genome-wide genotyping arrays or exome sequencing in the analysis of complete genetic variations. However, with a fixed budget, the high cost of sequencing many DNA samples is a limitation for GWAS (2–4). Recently, to reduce the cost of sequencing, a number of low (0.5-1x) or extremely low-coverage (0.1-0.5x) WGS has been carried out as an alternative method of genotyping (2, 5, 6). It is, however, unclear whether ultra-low coverage WGS (ulcWGS) below 0.1x data can capture enriched genetic variations across the entire allele frequency (AF) spectrum. When considering a balance between number of samples sequenced and sequencing read coverage, effective genotype imputation could provide more authentic DNA variants that would be helpful for genetic research.

Genotype imputation can be used to infer missing genotypes and to increase the accuracy of detecting genetic variants, such as single nucleotide polymorphisms (SNPs). In general, performance of genotype imputation is largely affected by sample size, sequencing coverage, analysis methods, and other parameters (7). A main challenge to use very low coverage WGS is how to achieve an adequately accurate imputation for downstream analyses. Previous attempts have shown the efficiency of low coverage WGS, for example, a high r^2^ of imputation accuracy observed by using 10 low coverage WGS (∼0.5x) as compared to known genotypes (6). Pasaniuc et al. reported that the GWAS signals obtained from using 909 whole-exome sequencing (∼0.24x) are comparable to using genotyping array (2). Gilly et al. found that more true association signals were identified by WGS (∼1.0x) than the traditional array-based study (5). Using ulcWGS (0.06x-0.1x) with 141,431 samples from a Chinese genomic study, the accuracy of imputed genotypes reached 0.71 (8). Even though the distribution of genetic background from large number of samples is expected to compensate for the low sequencing coverages, it has never been determined how many samples are needed to achieve a relatively high accuracy. More importantly, lack of comparative data with coverages less than 0.05x results in the limited application of ulcWGS to GWAS.

Gestational age is an important complex trait associated with biological processes and human disease. Biologically, gestational duration plays a vital role in both mental and physical health of children at an age of five-year-old (9). Gestational age shorter than 37 weeks is categorized as Preterm Birth (PTB). Previous studies found the contribution of both the maternal and fetal genomes to variation of gestational ages (10–12). However, they focused on European and African samples by involving few samples from Chinese or Eastern Asian ancestries. Overall, biological mechanisms underlying variation of gestational durations remain unclear, primarily because insufficient maternal or fetal genotypes with widespread gestational ages have been collected (13). Recently, Preimplantation Genetic Testing (PGT) with trophectoderm biopsy for embryo aneuploidy screening has become a common practice in *in vitro* fertilization (14, 15), and poses as an expectant source of genotypes for GWAS. However, if the average sequencing coverage of PGT is even lower than the lowest levels that have been reported in GWAS so far, it is necessary to examine whether such PGT datasets are appropriately applied to GWAS.

In this study, we devised a pipeline for analyzing and applying the ulcWGS of 17,844 embryo samples for GWAS. Our result shows that a large sample size is effective to increase the accuracy of genotype imputation even at an ultra-low coverage. Furthermore, using the imputed genotypes of 1,744 embryos that were successfully transferred and born with a widespread of gestational ages, we demonstrate the power of using ulcWGS in GWAS and provides insights into understanding genetic association of gestational age in embryos acquired from Chinese and East Asians. Refreshing the lowest coverage used in GWAS, our finding also provides a foundation for exploring the utilization of an even lower coverage for dissecting genotype-phenotype associations.

## Methods

### Samples and sequencing coverage

The whole PGT dataset of 17,844 embryos was from the Clinical Research Center for Reproduction and Genetics in Hunan Province, Reproductive and Genetic Hospital of China International Trust Investment Corporation - Xiangya. The protocol of embryo culture and biopsy was published in a related study (16). Three WGA kits were applied to the biopsied TE cells by following the manufactures’ guides, including REPLI-g Mini Kit (called MDA), WGA4 GenomePlex Single Cell Whole Genome Amplification Kit (called dop-PCR) and Rubicon Genomics PicoPlex Single Cell Whole Genome Amplification Kit (called PicoPlex). A 1-2 µg of the WGA product was subjected to library construction and sequencing on the four platforms, including BGI-Seq 500, Illumina MiSeq, Ion Proton, Ion Torrent (Additional file 1: Table S2).

### Study design

We developed a three-step pipeline to carry out genotype imputation using ulcWGS data and to perform GWAS of the detected SNPs (Fig. 1). Firstly, the raw reads of the 17,844 embryo samples were aligned to the hs37d5 reference genome. Sequencing coverage of these embryo samples displays a distribution with an average coverage 0.04x (Additional file 3: Figure S1). To our best knowledge, it is below the coverage of any dataset used previously for genotype imputation. After removing potential PCR duplicates, the aligned reads were used to call population SNPs. Secondly, we conducted genotype imputation on each individual sample at the called population SNPs and assessed the genotyping accuracy based on the standard Chinese sample HG005 with known genotypes from GIAB (Genome in a Bottle, NIST)(17). Last, we applied the imputed genotypes from 1,744 born embryos with complete follow-up records to GWAS and explored biological associations between the genetic variants detected in the born embryos and their gestational ages.

**Fig. 1.**
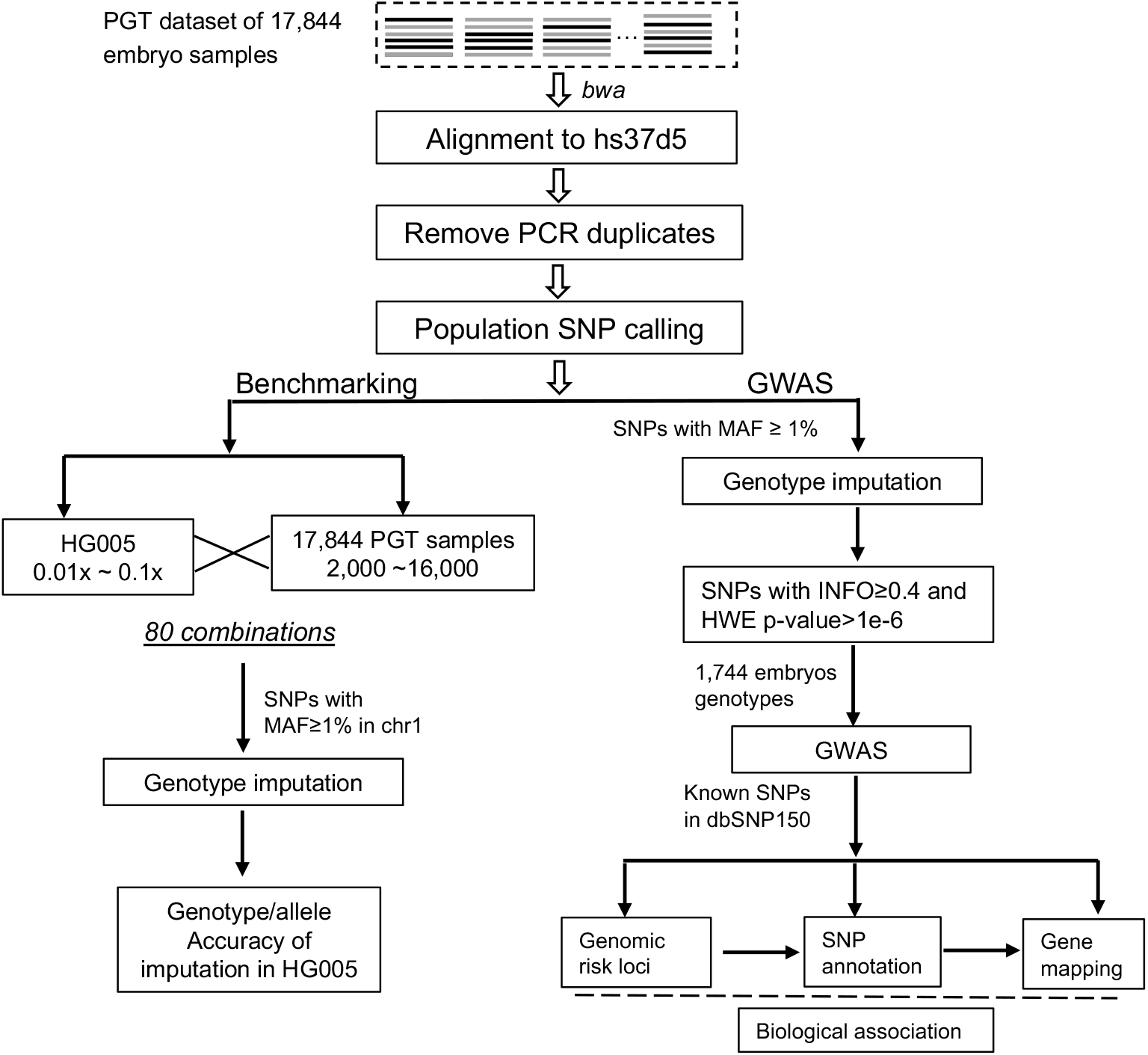
An overview of the analysis and benchmarking pipeline for ultra-low coverage WGS.

### Sequencing read processing and alignment

In the first step of Fig. 1, the raw reads of the PGT samples were delivered in two types, fastq or BAM. For BAM data, we used bedtools (18) to extract the raw reads into single-end fastq files. The raw reads of each sample were then aligned to the hs37d5 reference genome using BWA (19). BWA-mem was applied to samples sequenced by the Ion Torrent with longer reads and “bwa aln” was used for the rest of the samples with shorter reads. Samtools rmdup (20) was used to remove the potential PCR duplicates.

### Population SNP calling

In the population SNP calling stage, we modified the method of Liu et al. (8). The first stage is to use log-likelihood estimation for AF estimation, and the second stage is to use log-likelihood ratio test for determining allelic types. More details are described in Additional file 2: Supplementary Methods.

SNP calls of the raw population were filtered following the rules, 1) calls that overlapped with the 35-kmer problematic alignment regions in hs37d5 were removed (21); 2) calls that overlapped with regions with ENCODE mappability uniqueness score unequal to 1 were removed (22), tool “bigWigToBedGraph” used afterward to convert the bigwig into bed format) (23).

### Genotype imputation

We used STITCH (24) version 1.5.7 for genotype imputation. The ancestral haplotypes number *k* was set as 20, the assumed number of generations *nGen* was set to 2000, and the reads were binned into windows with *gridWindowSize* 10000. The *diploid* mode in STITCH was used. Although using a reference panel is optional in STITCH, we used the IMPUTE2 1000 genome haplotypes phase 1 reference panel as it improves the accuracy of imputation when the sample size is small. With sample size larger than 10,000, the improvement was not significant. All parameters were optimized by maximizing the *r*^2^ of the estimated AFs between imputation and population SNP calling in a randomly chosen 5 Mbp genomic region (chr3:180-185Mbp). When applying STITCH to the whole genome, we divided the genome into 5 Mbp windows with a 500 Kbp overlap between two windows.

For benchmarking, seqtk was used to subsample the HG005 Illumina WGS raw reads to 0.01x-0.1x (Paired-end 250bp, 300-fold). We aligned the reads to hs37d5 by using the same pipeline as used in the embryo samples. Because a computer takes a few years to impute whole genome with tens of thousands of samples, we worked on only chromosome 1, the longest one in human. The genotype imputation was benchmarked according to 80 combinations of the 10-scale coverages of HG005 with 8 sizes of our samples from 2,000 to 16,000, respectively.

All bi-allelic SNPs with MAF ≥ 0.01 found in chromosome 1 with the population SNP calling from the 17,844 samples are included for genotype imputation. We used the 167,814 SNPs both in our SNP callset and HG005 known genotypes for benchmarking. The imputed genotypes were compared to the truth released by GIAB for estimating the imputation accuracy. Then, genotype imputation was applied to all 17,844 embryo datasets at the 31,622,332 bi-allelic sites with MAF ≥ 0.01 found in population SNP calling. The entire imputation process spent 19 days and used 15 machines with 16 cores (two 8-core Intel Xeon Silver 4108 CPU). Two filters “INFO score ≥ 0.4 and HWE *p-*value >1e-6” were applied to select the imputed genotypes.

### Genome-wide association study (GWAS)

To conduct GWAS, we used score statistics (25) that is implemented in ANGSD (26). Variants satisfying four conditions were selected as inputs, including 1) known in dbSNP150, 2) MAF ≥ 0.01, 3) INFO score ≥ 0.4, and 4) HWE *p*-value > 1e-6. To remove biases, we specified 16 covariates for ANGSD, 8 most significant principal components calculated from the inputs of PCA (Additional file 2: Supplementary Methods), and 8 clinical records including maternal age, maternal BMI, fetal sex, either parent with single-gene disease, either parent with chromosome abnormality, multiple pregnancy, preeclampsia and gestational diabetes of mellitus. Except the default parameters, we set *minHigh* to 15 (requiring at least 15 high credible genotypes from the input) instead of 10 to achieve better accuracy with a large sample size. ANGSD did not create an output beta-coefficient, so we followed the ideas of Skotte et al. (25) by incorporating the genotype probabilities and all 16 covariates into a linear regression model, with the gestational age as a response variable. The effect size was calculated by the coefficient of genotypes.

### Independent SNPs and genomic risk loci

The significant SNPs from our GWAS were mapped to genomic risk loci using FUMA pipeline (27) and the LD information of 1000G EAS variants (28). We first defined “independent significant SNP”, a SNP that meets genome-wide significance level (p value ≤ 4.515e − 8) and is independent of other significant SNPs (with LD r^2^ < 0.6). FUMA also generated a set of lead SNPs with low LD and with other (r^2^ < 0.1) from the independent significant SNPs. The genomic risk loci were identified by starting from these lead SNPs and through iteratively merging related genomic regions to them according to FUMA’s rules.

Also, the FUMA pipeline sorted out a set of candidate SNPs from our inputs that meets one of two conditions, 1) the independent significant SNPs and 2) SNPs that are linked to the independent significant SNPs (with LD r^2^ ≥ 0.6). For condition 2, the SNPs can be from our imputed genotypes if p value below 0.05 or from the reference panel of 1000G EAS. ANNOVAR was used to annotate the candidate SNPs (29).

### Functional annotation of the mapped genes

DAVID online tool (30) was used to analyze the enrichment of Gene Ontology (GO) biological processes and KEGG pathways for the coding genes mapped to the risk loci.

### Gene mapping

We used three gene-mapping strategies provided by FUMA (27), including positional, expression quantitative trait locus (eQTL) and chromatin interaction. For positional mapping, ANNOVAR annotations were used. The candidate SNPs were mapped to the nearest genes within a maximum 10 Kbp distance. For eQTL mapping, expression data of all tissue types in GTEv6, GTEv7, and GTEv8 (31) were used. We required False Discovery Rate (FDR) < 0.05 and p value <0.001 for a valid eQTL mapping. All chromatin interaction data in FUMA were used (32–35). The promoter was set to upstream 2000 bp to downstream 500 bp of transcriptional starting sites. We required FDR < 1e-6 for a valid chromatin interaction mapping.

### Analysis of genome-wide mRNA expression data

We first extracted the genome-wide microarray and RNA-seq data of human mRNA expression from GEO/NCBI database. The mRNA data includes three subsets in maternal PTB, infant PTB and breast cancer (Additional file 1: Table S8). Based on the normalized expression data provided by the database, we analyzed differentially expressed genes (DEGs) between different conditions, including 1) PTB vs. normal term, 2) BPD or sepsis vs. infant without BPD or sepsis, and 3) breast cancer vs. control samples. For microarray platform-based data, we used the *limma* package in R programming language and conducted empirical Bayes moderated t-test. DEGs were detected with a fold change above 1.5 and p value below 0.05. For RNA-seq data with raw counts, we utilized the Edge-R method to identify DEGs. The DEGs are listed in Additional file 1: Table S9.

## Results

### Benchmarking genotype imputation using the ultra-low coverage sequencing data of 17,844 embryos and HG005

We estimated the accuracy of the imputed genotypes from the SNPs of chromosome 1 both called in our 17,844 samples and the known genotypes in HG005. The genotype imputation was benchmarked according to 80 combinations of the 10-scale coverages of HG005 with 8 sizes of our samples. A monotonic increase in accuracy with sequencing coverage was observed (Fig. 2a), consistent with previous studies (2, 24). The accuracy for sample size of 2,000 stayed at around 0.48 under all sequencing coverages. But for a larger sample size of 16,000, its accuracy increased from 0.48 at 0.01x to 0.66 at 0.1x. This result suggests that at ultra-low coverages, increase in sample size could obtain higher accuracy (Additional file 1: Table S1). In general, a lower coverage with a larger sample size results in better performance than a higher coverage with a smaller sample size. For example, the genotype accuracy at 0.05x with 14,000 samples versus 0.1x with 4,000 embryos was 0.61 versus 0.55. A larger sample number is therefore more efficient in optimizing genotype imputation than increasing sequence coverage. It is also noticed that at the two lowest coverages in our experiments, the contribution of increasing sample size was not significant and the accuracy plateaued at 0.52 (0.01x) and 0.55 (0.02x). Using the same datasets, we evaluated allele accuracy that relaxed zygosity correctness from genotype accuracy. The corresponding accuracies were much better (increased to 0.7 and higher) while maintaining the same trend with increasing sample size and coverage (Fig. 2b). Therefore, when the genotypes are incorrectly imputed for some SNPs, the non-reference allele could be correctly detected.

**Fig. 2.**
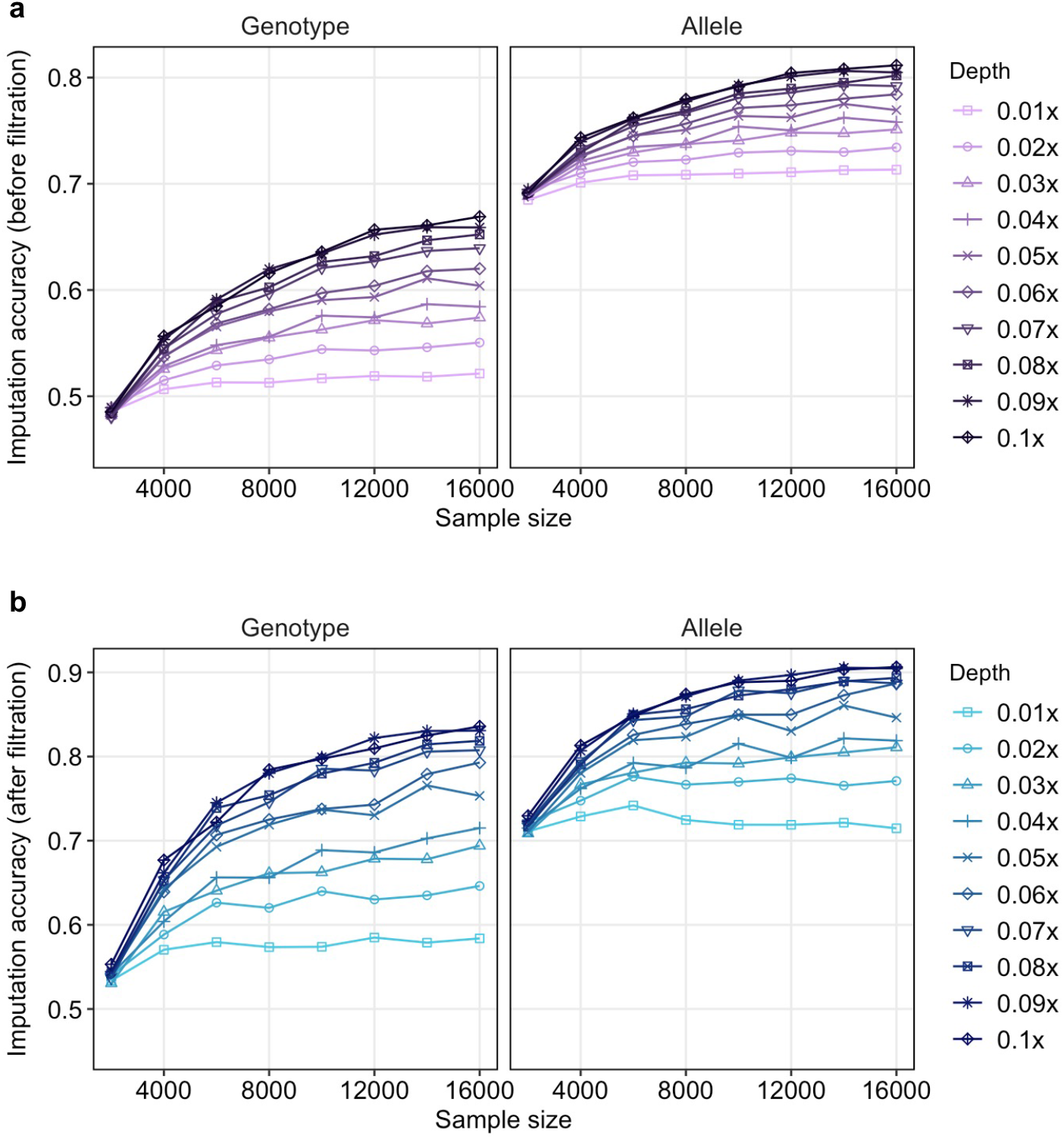
Imputation accuracy at different coverages and sample sizes. The accuracies of imputed genotype (a) or allele (b) were obtained by comparing with the known genotypes in HG005. After using filter “INFO score ≥ 0.4 and HWE *p*-value > 1e-6” the accuracies of imputed genotype (c) or allele (d).

We next examined several important quality metrics that are widely used to filter falsely imputed genotypes. The INFO score of IMPUTE2 style (36) denotes the certainty of an imputed genotype and has been accepted as a quality metric of imputation. For a combination of 0.04x coverage with 16,000 samples, we benchmarked ten different INFO score thresholds from 0.1 to 1.0 and detected corresponding SNPs. As increase in the INFO scores, we observed a consistent increase in genotype accuracy from ∼0.60 to 0.99, but a rapid decrease in number of SNPs that meet these thresholds (Fig. 3a). Thus, INFO score could act as an effective metric to evaluate the accuracies at ultra-low coverage, but its thresholds should be meticulously chosen in order to retain sufficient SNPs. Effect of MAF scores on genotype accuracy of HG005 were then tested as a potential metric. We divided the genotyping results at different sequencing coverages (sample size fixed to 16,000) into bins of MAF ranges (0.01 MAF a bin) and calculated the genotype accuracy each bin. The genotype accuracy increased rapidly from MAF 0 to 0.05 and reached a turning point at 0.05. After this point, the accuracy became slow increasing (Fig. 3b). However, even for the most common SNPs (MAF 0.4∼0.5), the accuracy was converged at ∼70%. The accuracy of the two lowest coverages 0.01x and 0.02x fluctuated especially at low MAF cutoffs. Because such fluctuation was not observed during INFO testing, MAF might not be a reliable metric to change genotype accuracy at ultra-low coverage. In subsequent analyses, we followed the common practice to use SNPs with MAF ≥ 0.01 for GWAS. Finally, we combined HWE *p*-values with INFO scores as a filter but without losing too many SNPs. HWE *p*-values could evaluate the probability of the imputed genotype at a certain SNP that is significantly different from the expectations by Hardy-Weinberg Equilibrium. We summarized the genotype accuracy of different combinations of INFO scores and HWE *p*-value cutoffs in Table 1. When INFO scores were set above 0.4, the accuracies of genotype and allele were 70.0% and 83.4%, respectively, with 48,176 SNPs left. The “INFO score ≥ 0.4 and HWE *p*-value >1e-6” resulted in an increased accuracy 71.5%, with 28773 SNPs left. Thus, our GWAS utilized this setting, “INFO score ≥ 0.4 and HWE *p*-value >1e-6” as filtering criteria.

**Fig. 3.**
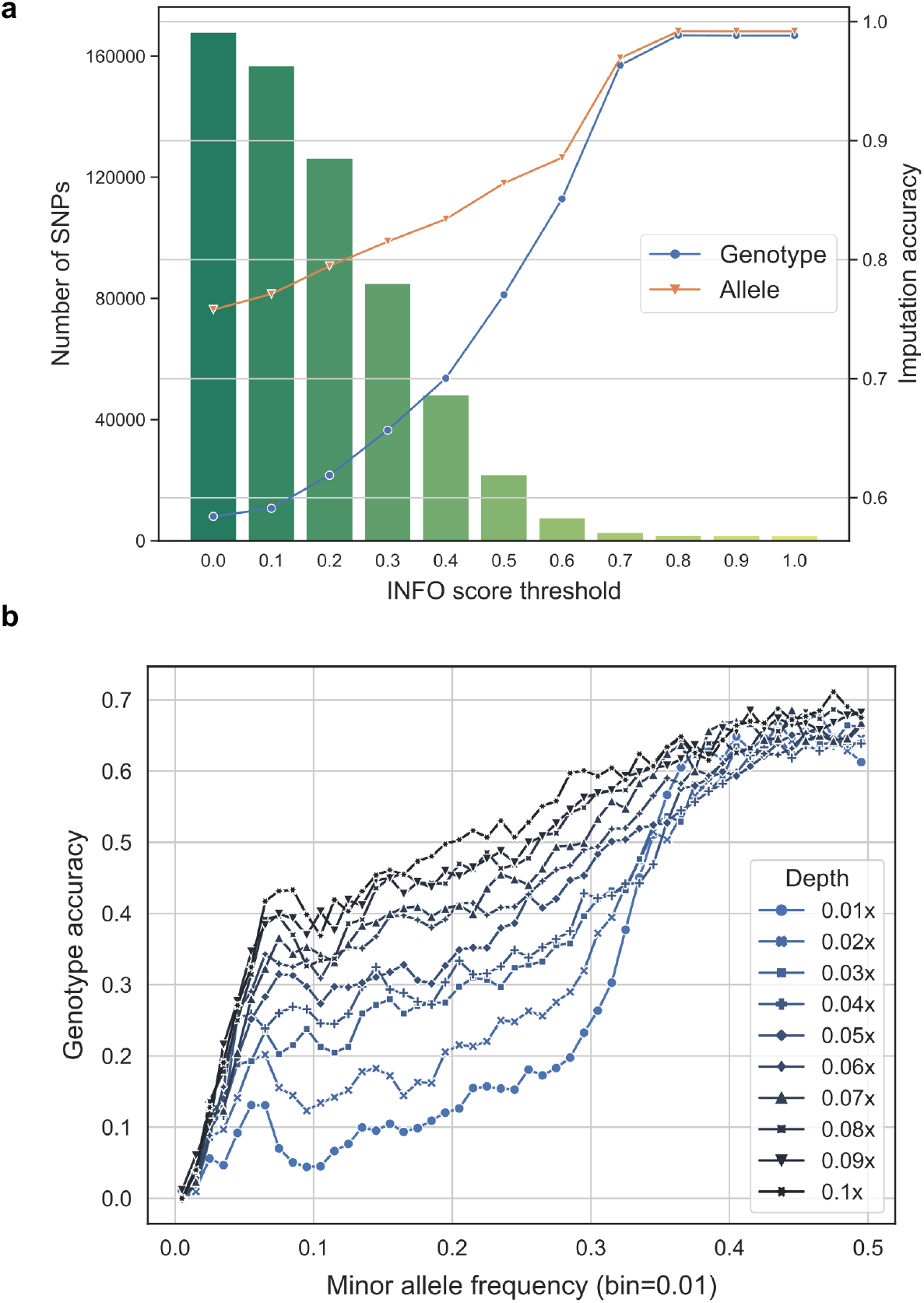
Performance of different imputation result filters. The accuracies of our samples were calculated against the known genotypes in HG005. a, effect of INFO score filtering cutoffs on genotype and allele accuracies. The imputation was conducted by using 0.04x sequencing coverage of HG005 and with 16000 embryo samples. b, effect of MAF cutoffs on genotype accuracy at multiple sequencing coverages. The imputation was conducted through different sequencing coverages of HG005 with 16000 embryo samples.

**Table 1.**
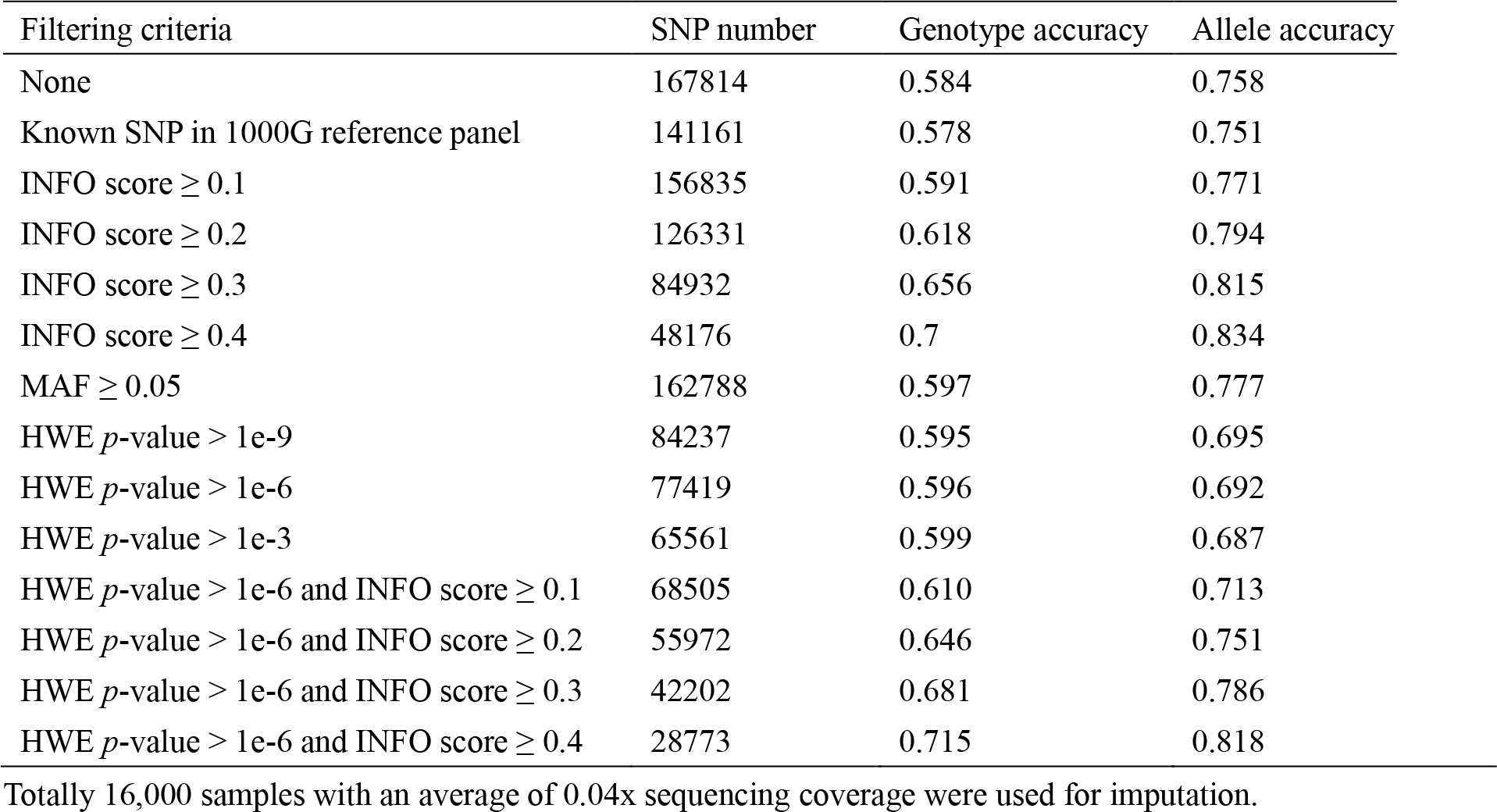
Genotype imputation performance with different filtering criteria.

| Filtering criteria | SNP number | Genotype accuracy | Allele accuracy |
| --- | --- | --- | --- |
| None | 167814 | 0.584 | 0.758 |
| Known SNP in 1000G reference panel | 141161 | 0.578 | 0.751 |
| INFO score $\geq 0.1$ | 156835 | 0.591 | 0.771 |
| INFO score $\geq 0.2$ | 126331 | 0.618 | 0.794 |
| INFO score $\geq 0.3$ | 84932 | 0.656 | 0.815 |
| INFO score $\geq 0.4$ | 48176 | 0.7 | 0.834 |
| MAF $\geq 0.05$ | 162788 | 0.597 | 0.777 |
| HWE $p$-value $> 1e-9$ | 84237 | 0.595 | 0.695 |
| HWE $p$-value $> 1e-6$ | 77419 | 0.596 | 0.692 |
| HWE $p$-value $> 1e-3$ | 65561 | 0.599 | 0.687 |
| HWE $p$-value $> 1e-6$ and INFO score $\geq 0.1$ | 68505 | 0.610 | 0.713 |
| HWE $p$-value $> 1e-6$ and INFO score $\geq 0.2$ | 55972 | 0.646 | 0.751 |
| HWE $p$-value $> 1e-6$ and INFO score $\geq 0.3$ | 42202 | 0.681 | 0.786 |
| HWE $p$-value $> 1e-6$ and INFO score $\geq 0.4$ | 28773 | 0.715 | 0.818 |

Totally 16,000 samples with an average of 0.04x sequencing coverage were used for imputation.

To summarize our benchmarking results for future study on ulcWGS, we built a regression model (Formula 1) to calculate the expected genotype accuracy using sequencing coverage and sample size as inputs.

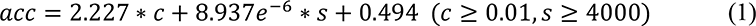

where *acc* is the expected genotype accuracy, *c* denotes the sequencing coverage, *s* denotes the sample size. The model has a r^2^ of 0.874 (Additional file 3: Figure S2).

### GWAS of gestational ages using 1,744 born embryos

With the solid foundation laid out in the previous section, we have obtained sufficient good quality SNPs for GWAS. Among the 17,844 sequenced embryos, 1,744 were transferred and gave birth to a baby. The gestational age of all 1,744 born embryos are well documented and thus were chosen for biological associated study. We revised the population SNP calling method used in Liu et al. (8). A total of 151,793,444 SNPs were detected and of 141,718,305 are bi-allelic. The MAF spectrum bi-allelic novel and known variants in dbSNPv150 (37), 1000G (28) and gnomAD (38) is shown in Additional file 3: Figure S3a. The transitions/transversions ratio for “all bi-allelic SNPs” and “bi-allelic SNPs known in dbSNPv150” were 3.05 and 3.58, respectively. Pearson correlation coefficient of the non-reference AFs between the 301 Chinese samples in 1,000G (so-called 1000G CHN (28)) and the corresponding SNPs obtained in our dataset was 0.986 (Additional file 3: Figure S3b). This result supports a strong correlation between the two datasets, and high confidence of the known variants used in our analysis. We also performed genotype imputation at bi-allelic population SNPs with MAF ≥ 0.01. Pearson correlation coefficient was 0.985, showing a high consistency between the estimated AFs in population SNP calling and in imputation.

Three different whole genome amplification (WGA) methods and four different sequencing platforms were used in the PGT dataset (Additional file 1: Table S2). It is not uncommon that large number of samples may use multiple sequencing platforms and WGA. Removing these unrelated covariates from GWAS as much as possible is essential especially when the sequencing coverage is ultra-low. Such covariates should be detected and disregarded in GWAS. We applied Principal Component Analysis (PCA) to the imputed genotypes of SNPs with MAF ≥ 0.05 among all 17,844 embryo samples (Additional file 2: Supplementary Methods, Additional file 3: Figure S4a). The first and second principal components distinguish the differences of sequencing platforms (Additional file 3: Figure S4b) and of WGA methods (Additional file 3: Figure S4c). Therefore, we used the top eight principal components and eight other clinical records as covariates in GWAS. PCA was also applied to the GWAS samples and the top principal components were included in the subsequent analyses as covariates for removing the biases.

We used the state-of-the-art one-stage GWAS strategy (39) to analyze the 1,107,198 imputed SNPs in the 1,744 transferred and born embryo samples with complete follow-up records. The distribution of gestational ages shown in Additional file 3: Figure S5 include 162 preterm deliveries (gestational age < 37 weeks), 42 early preterm deliveries (gestational age <34 weeks), and 8 very early preterm deliveries (gestational age <28 weeks). The gestational ages were standardized by z-score and incorporated as a quantitative trait.

A total 1,107, 198 SNPs with imputed genotypes were selected for GWAS that are in accord with, 1) MAF ≥ 0.01, 2) known in dbSNPv150, and 3) passed the filter “INFO score ≥ 0.4 and HWE *p*-value >1e-6”. The Q-Q plot shows a large deviation of the observed p values from the null hypothesis (Additional file 3: Figure S6). The linkage disequilibrium score regression (LDSC) software package (40) with 1000G EAS reference was used to estimate *λ_GC_* = 0.992, mean *χ*^2^ = 1.012. The LD score regression intercept was 0.952, standard error = 0.021, indicating that the population stratification and other factors were well-controlled. We identified 40 significant SNPs satisfying Bonferroni-corrected significant levels of 4.515e-8. The Manhattan plot shows the distribution of the detected SNPs cross all chromosomes (Fig. 4a, Additional file 1: Table S3).

**Fig. 4.**
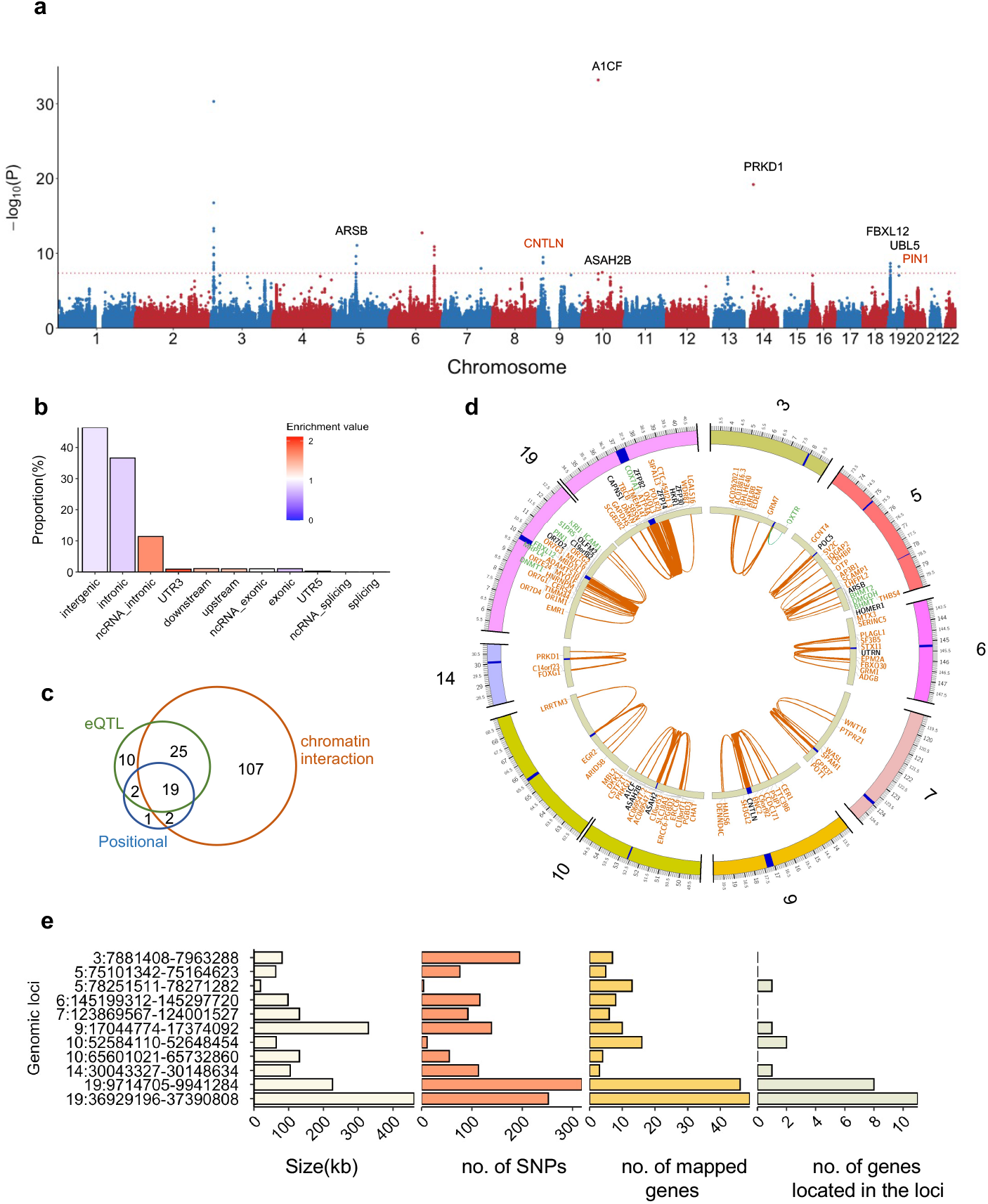
SNP-based genome-wide association on gestational age. a, Manhattan plot of the SNPs in GWAS. The red dash line represents the genome-wide significance level 4.515e-8. The SNP “rs946934582” with p value of 2.764e-144 is beyond the scale, thus hereby listed alone. The genes shown are linking with the candidate SNPs and position of the corresponding genomic risk loci. b, functional annotation and enrichment test result of the candidate SNPs in FUMA. c, a Venn diagram of the 166 genes that could be mapped to the 11 genomic risk loci by positional, eQTL and chromatin interaction strategies. d, a Circos plot of the chromatin interactions and eQTL mapping in the 11 genomic risk loci from eight chromosomes. The outer ring is chromosomes, the regions in blue denote genomic risk loci. The middle ring represents the mapped genes. The color of the gene symbols shows how they were mapped, eQTL in green, chromatin interaction in orange, and both eQTL and chromatin interaction in black. The inner ring shows the linking edges, eQTL in green, and chromatin interaction in orange. e, a summary of the 11 genomic risk loci.

We used FUMA (27) pipeline for GWAS downstream analysis. First, FUMA generated a set of candidate SNPs and the 11 independent SNPs (Additional file 1: Table S4). By annotation, most of the SNPs are located and enriched in intergenic and intronic regions (Fig. 4b), which is similar to the previous study (41). Specifically, there are 11 SNPs located in exons (0.8% of total), and 4 of them are non-synonymous SNPs (Additional file 1: Table S5). Additionally, we observed the distribution of regulatory elements and chromatin states with the candidate SNPs (Additional file 3: Figure S7). According to RegulomeDB scores assigned to each candidate SNP, 1.09% SNPs were classified as likely to affect regulator binding (score 2a and 2b) and 0.21% as likely to affect regulator binding and linked to expression of a gene target (score 1d and 1f). These proportions of SNPs hold a relatively high likelihood to affect the regulatory elements along noncoding regions. Second, we identified 11 leading SNPs from their corresponding genomic risk loci by FUMA. Fig. 4e shows the DNA length, number of SNPs and mapped genes of these risk loci. The zoom in locus plot of the 11 risk loci are shown in Additional file 3: Figure S8. Of 4 risk loci were reported to be associated with refractive astigmatism, adolescent idiopathic scoliosis, glomerular filtration rate, among others, indicating a possible connection of these diseases or traits with PTB (Additional file 1: Table S6).

By integrating strategies positional, eQTL and chromatin interaction mappings, we identified a total of 166 genes mapped to the 11 risk loci, including 48 and 19 genes from two and three strategies, respectively (Fig. 4c, Additional file 1: Table S7). There were 24 genes detected within or less than 10 Kbp from the candidate SNPs and 7 of them were shown in Fig. 4a based on the location of the genomic risk loci. Importantly, *CNTLN* was reported as a PTB related gene (42), and *PIN1* involves inhibition of breast cancer (43). A Circos plot shows the graphic distribution of the mapped genes via eQTL and chromatin interaction, and their links with the genomic risk loci (Fig. 4d). The breakdown of each chromosome is shown in Additional file 3: Figure S9. Even though not within any risk loci, *CRHBP* was linked through chromatin interaction mapping to two loci, chr5:75101342-75164623 and chr5:78251511-78271282. Enrichment analysis of DEGs in 30 tissue types in GTEx v8 (44) exhibits significant overexpression of the mapped genes in both ovary and uterus (Additional file 3: Figure S10a). The gene-set enrichment analysis also indicates their association with the immune system, breast cancer and transcriptional regulation (Additional file 3: Figure S10b).

### Association of the 166 mapped genes from GWAS with preterm birth, infant disease and breast cancer

GWAS of gestational age related PTB has been implicated in biological functions that include immune response, inflammatory response, and coagulation factors (11, 45–47). We compared our 166 mapped genes with reported PTB markers by collection of 8 published resources, here classed to 3 PTB sets, including dbPTB from Sheikh et al. (48) and Uzun et al (49), PTB-merged from 5 data resources (10), and PNAS-identified DEGs of PTB in 2019 (10). We found that *CRHBP, EMR1, ICAM1, MBL2, OXTR*, and *THBS4* have been reported in at least 2 PTB sets. Specifically, *ICAM1* was present in all 3 sets and 6 data resources, and both *CRHBP* and *OXTR* in 5 resources (Additional file 1: Table S7). In addition, there were 8 genes overlapped with 1 PTB set (Fig. 5a). This result pinpoints a relationship of the detected risk loci with PTB, and possible roles of the overlapped genes in PTB. There are totally 1,930 genes reported as PTB-related; however, only 50 were frequently recognized by at least 5 data resources, hereafter referred to as PTB marker genes (Additional file 1: Table S7). The 50-PTB marker set was significantly enriched with inflammatory and immune response related processes or pathways (Fig. 5b). Similarly, those PTB genes that overlapped with 3 or 4 resources mainly participate in the same biological functions. The PTB-related genes listed at Fig. 5a involve immune response (*EMR1, ICAM1, PTPRZ1* and *MBL2*), inflammatory response (*CRHBP, ICAM1, PTPRZ1* and *MBL2*), coagulation (*MBL2*), apoptosis (*PLAGL1*) and cell adhesion (*ICAM1* and *THBS4*), emphasizing their associations with PTB.

**Fig. 5.**
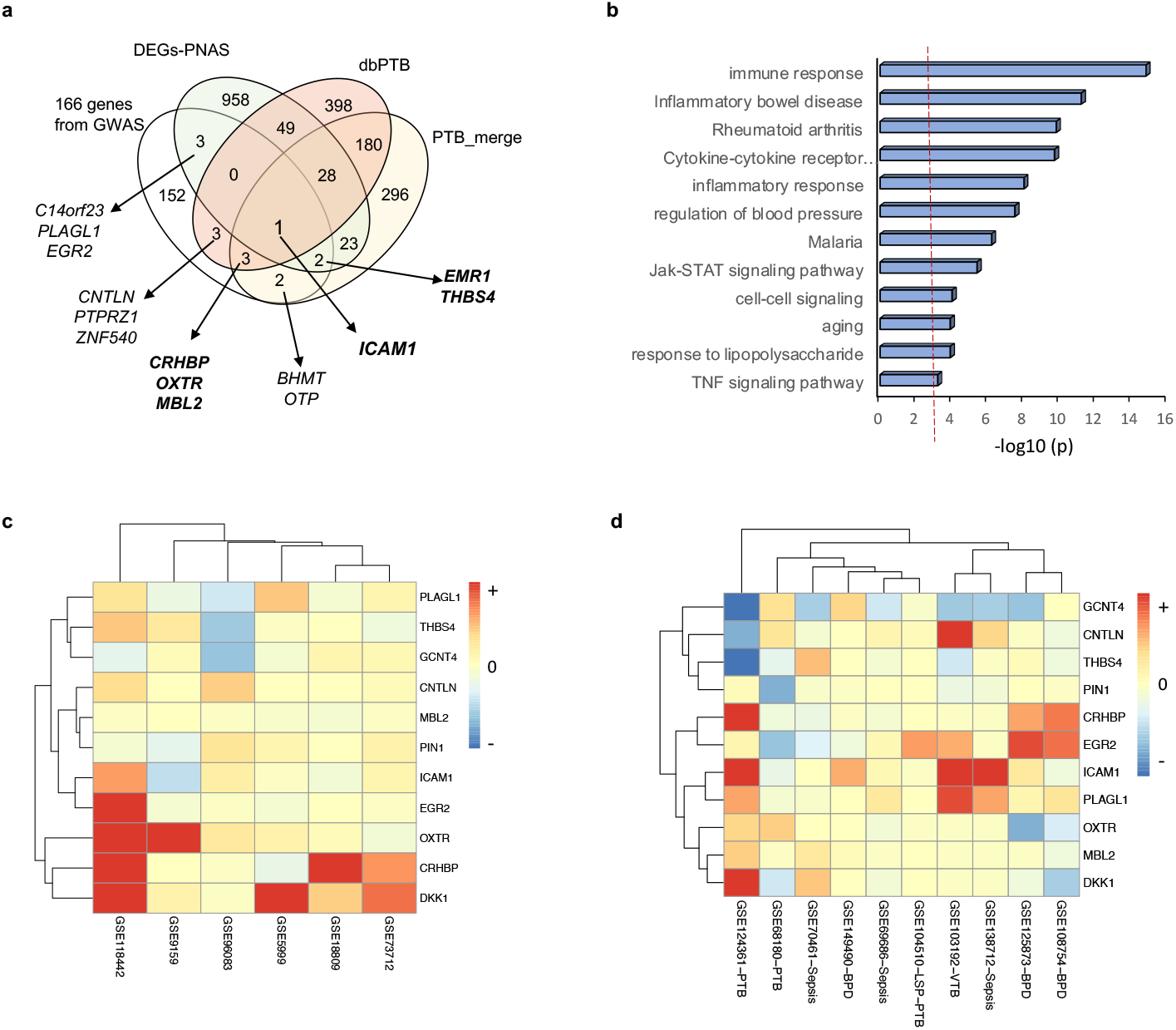
Comparison of the 166 genes mapped to 11 genomic risk loci with PTB and infant disease. a, the 166 mapped genes were compared with 3 sets of reported PTB genes including dbPTB from Sheikh et al. (48) and Uzun et al (49), PTB-merged from 5 resources (10), and PNAS-identified DEGs in 2019 (10) (see Additional file 1: Table S8). b, a bar graph showing significantly enriched GO biological processes and KEGG pathways based on the 50 PTB marker genes (see Additional file 1: Table S7). c-d, heatmaps showing expression profiling and clusters of the PTB genes predicted from GWAS under maternal PTB (c) and PTB infant with BPD and sepsis (d). The gene expression data was extracted from the listed GSE accession numbers of NCBI/GEO.

To determine the relationship between the mapped genes and PTB, we first analyzed the DEGs between maternal PTB and normal term birth based on six published datasets of genome-wide gene expression (Additional file 1: Tables S8 and S9). Significant overexpression of *CRHBP* and *OXTR* were identified in two datasets, while overexpressed (*ICAM1, EGR2*, and *PLAGL1*) and underexpressed (*THBS4*) genes were present in one dataset (Table 2). Another *DKK1* expressed higher levels above 2.0-fold in two datasets, suggesting its importance in PTB. Then, we analyzed four infant PTB datasets of gene expression, and found differentially increased expression of *ICAM1, CRHBP, DKK1, EGR2*, and *PLAGL1*, consistent with maternal PTB. In contrast, significantly underexpression of *THBS4, PIN1* and *GCNT4* was commonly detected by maternal and infant subgroups (Table 2). It is also noticed that *DNMT1* shows increased expression in both fetal and maternal groups (Table 2). DNA methylation was suggested to involve generation of early PTB (10); however, the role of *DNMT1* as a DNA methyltransferase in PTB has not been determined yet. This analysis provides evidence that these mapped DEGs above including *DNMT1* are associated with both maternal and infant PTB. Relatively, *OXTR* seems to be related to only maternal factor. A heatmap of mRNA expression shows clustering of 6 expression profile cross PTB and normal term conditions (Fig. 5c). Two clusters display distinct expression patterns of these PTB genes.

**Table 2.**
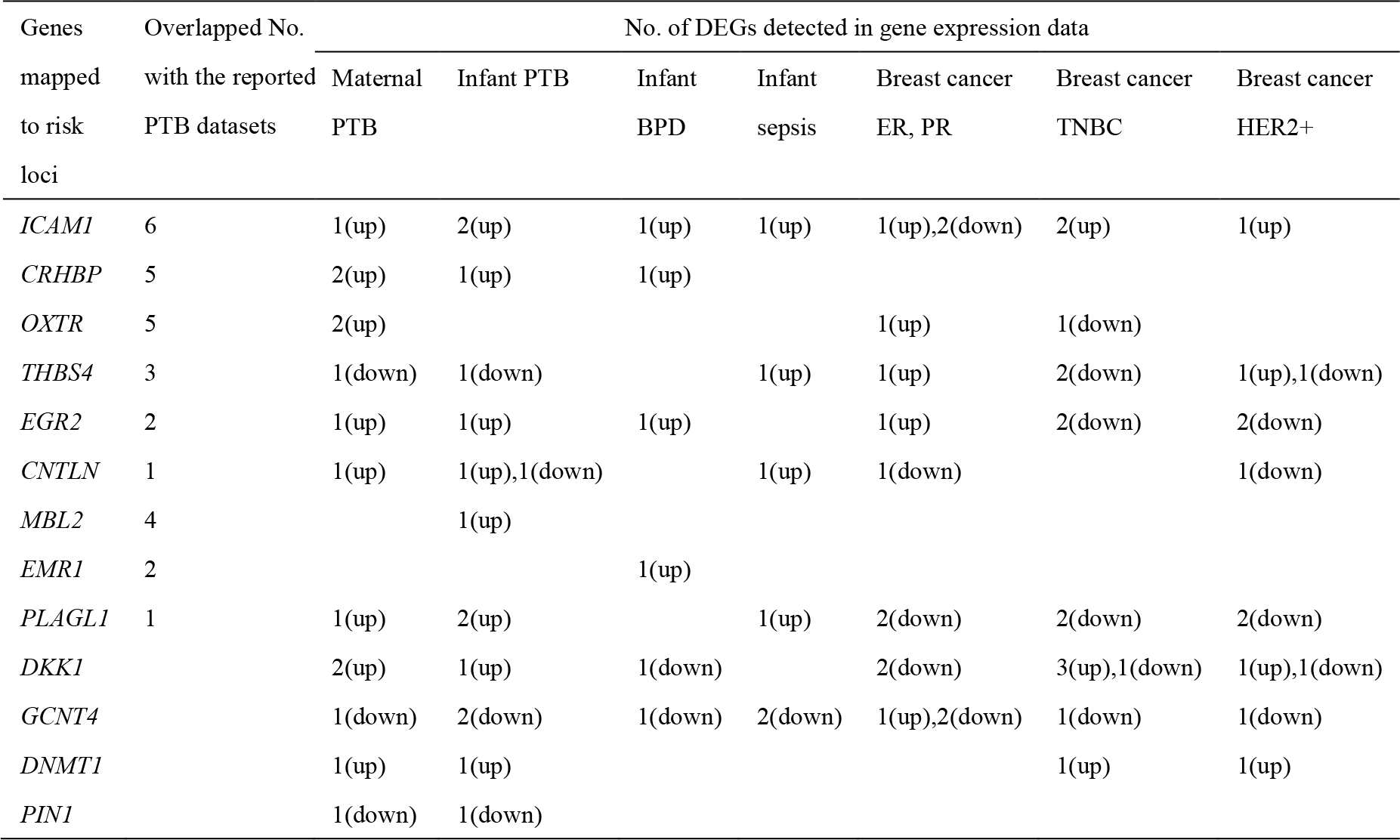
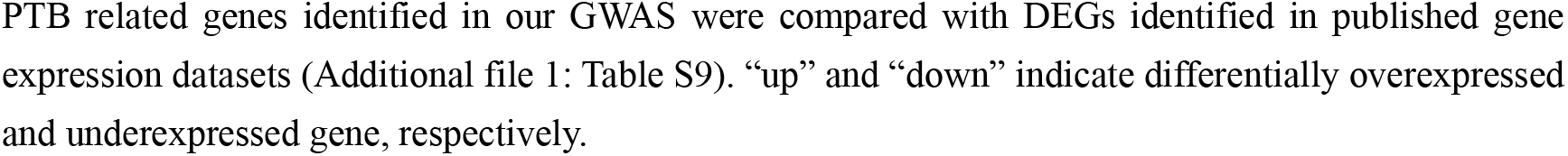
Main mapped genes differentially expressed in PTB, infant disease and breast cancer.

It is well known that bronchopulmonary dysplasia (BPD) is the most common respiratory disorder among children born preterm (50, 51). The pathogenesis of BPD involves multiple prenatal and postnatal mechanisms affecting the development of immature lung. Also, neonatal sepsis is associated with severe morbidity and mortality during the neonatal stage. The incidence of late-onset sepsis increases with increase in survival rate of preterm and low weight babies (52). Thus, we examined the possible relationship of the 166 genes with infant BPD and sepsis by analyzing gene expression data derived from samples of preterm infants (Additional file 1: Table S8). We first identified differentially increased expression of *CRHBP, ICAM1* and *EGR2* under PTB of maternal & infant and BPD conditions, but differentially decreased expression of *DKK1* (Table 2). Under sepsis condition, differentially overexpressed *CNTLN, ICAM1*, and *PLAGL1* in PTB were consistently observed. By contrast, *GCNT4* expression was always significantly decreased under maternal and infant subpopulations. The gene clustering shows diversity of gene expression across different samples with infant PTB; however, these up-regulated genes were grouped together (Fig. 5d), supporting the idea that BPD or sepsis induces the change in these PTB genes.

Breast cancer is one of the most frequently diagnosed malignancies observed during pregnancy. It often presents characteristics of high malignancy and are hormone receptor negative like Estrogen receptor (ER)-, HER2+ or triple negative breast cancer (TNBC). We collected gene expression data mainly presenting three subtypes of breast cancer, TNBC, HER2+ and ER or Progesterone receptor (PR) (Additional file 1: Table S8). By analysis of DEGs, we detected the expression of PTB related genes *DKK1, ICAM1, DKK1, EGR2, PLAGL1, GCNT4* and *THBS4* in all three subtypes. Increased *ICAM1* and decreased *PLAGL1* were consistently identified in these datasets (Table 2). In fact, *ICAM1* has been reported as TNBC markers (53) and acts as prognostic molecule of breast cancer (54). Both *ICAM1* and *DKK1* could increase expression in TNBC cells (55). However, other PTB related genes do not display similar changes in expression under the cancer subtypes. For example, *OXTR* was detected by only one dataset of ER+/-, while *CRHBP, EMR1, PIN1* and *MBL2* were not found among any of the subtypes. Conversely, underexpression of *PLAGL1* and *GCNT4* were found in all three types of breast cancer. In addition, overexpression of *DNMT1* was observed in TNBC and HER2+ subtypes, that validates its oncogenic roles in breast cancer and drug target of TNBC (56, 57).

To identify further interactions between the selected PTB genes from Table 2 and the top 50 PTB markers, we calculated Pearson correlation coefficient by comparing their gene expression of maternal and infant PTB, BPD and sepsis subsets, respectively. We built the corresponding co-expression networks of the PTB genes (Additional file 3: Figure S11). Clearly, the co-expressed genes involve immune and inflammatory responses, coagulation, as well as apoptosis, angiogenesis, among others. We then constructed the networks cross different subpopulations. As shown in Fig. 6a, the PTB genes could involve both maternal and infant PTB processes, especially *ICAM1*, *PLAGL1, EGR2* and *CRHBP* that link to TLR4, a known preterm marker associated with immune and inflammatory processes (58). Similarly, co-expression interactions of these genes with the top PTB markers were observed under maternal PTB and infant BPD (Fig. 6b). Both *OXTR* and *PIN1* only display gene correlations under maternal PTB, whereas *MBL2* only correlates under infant PTB. This analysis indicates that the predicted PTB-related genes including *ICAM1*, *CRHBP*, *PLAGL1, EGR2, CNTLN*, and *DKK1*, play an important role in preforming biological activities associated with PTB, infant disease, possibly breast cancer, due to the gestational age-induced.

**Fig. 6.**
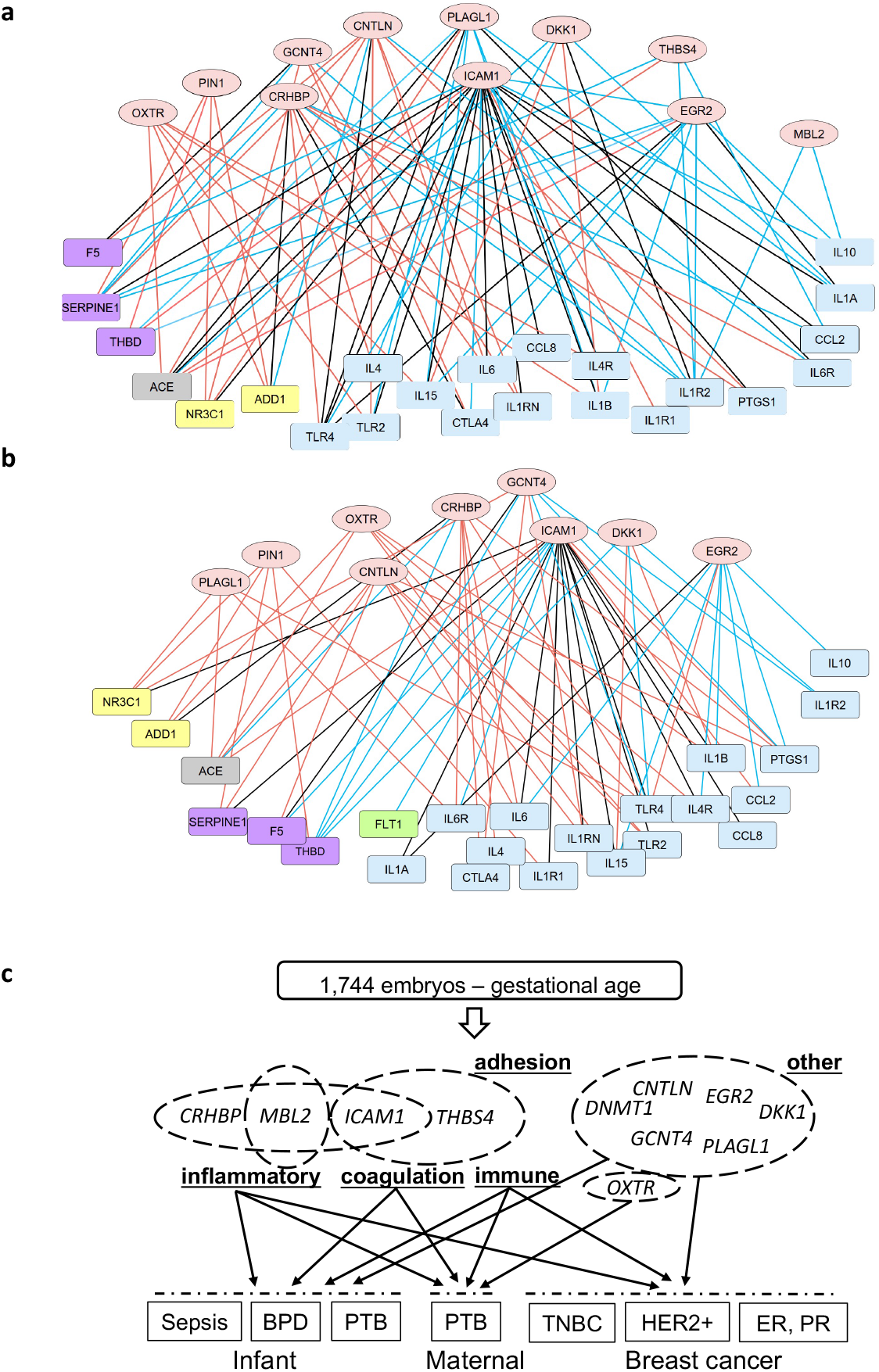
Associated analysis of the PTB-related genes in maternal and infant subtypes. a, a gene co-expression network merging maternal and infant PTB subtypes. b, a gene co-expression network merging maternal PTB and BPD of preterm infant. In A and B, oval nodes represent PTB related genes predicted from GWAS using the 1,744 born embryo samples, rectangle nodes refer to the 50 top PTB markers summarized from previously studies (Additional file 1: Table S7), involving immune or inflammation (blue), apoptosis (yellow), angiogenesis (green), coagulation (purple) and other biological processes (grey). Co-expressive edges (Pearson correlation p value < 0.01) linking nodes represent maternal (red), infant (blue) and both maternal and infant (black). c, a graphic summary to illustrate gestational age’s association with PTB, infant disease and breast cancer.

## Discussion

Low or very low coverage sequencing data have been increasingly used in discovery of genetic variation and GWAS. However, if the coverages are further decreased to extremely ultra-low levels, how many samples are required to obtain a relatively high accuracy of genotype, and whether these samples can be appropriately applied to GWAS. To address this challenge, we used 17,844 PGT datasets of embryo samples with an average of 0.04x coverage and achieved genotype imputation performance comparable to those using low coverage samples. We first demonstrate that increase in number of ulcWGS samples is more efficient than changing sequencing coverages and indicates the ability of such ultra-low coverage PGT samples to obtain adequate accuracy of phenotypes. Furthermore, we found that INFO score and HWE *p*-value are effective to act as filters to improve the accuracy at ultra-low coverages while meanwhile keep enough SNPs for downstream analyses. To our best knowledge, the samples used in this study hold the lowest average WGS coverage for GWAS so far, and our study provides a framework for guiding other researchers who work on ulcWGS data.

Gestational age is a multi-factor phenotype involved in maternal and fetal biological activities (59). To investigate effects of the fetal genome on gestational age, we used the imputed genotypes of 1,744 born embryo samples, a cohort of samples who successfully gave birth after *in vitro* fertilization. From the 166 genes mapped to the 11 genomic risk loci, we identified a set of the PTB-related genes that were previously reported (12, 59). *CRHBP, ICAM1*, and *OXTR* are primary representative genes showing evidence of genetic association with gestational age among our samples. *CRHBP* is an important gene in maternal and fetal gestation that could regulate the pregnancy length by increasing/decreasing the concentration of CRH (60, 61). *ICAM1* involves disease induced PTB during pregnancy (62–64). Oxytocin signaling is mediated by Oxytocin receptor (*OXTR*), which is related to gestational age (65). We validated these predicted PTB genes by analysis of DEGs from maternal and infant PTB subpopulations.

PTB is neonatal birth occurring before 37 weeks of gestation age and is a leading cause of infant morbidity and mortality. Understanding of genetic and molecular mechanisms of PTB and its association with gestation duration is currently insufficient. Recently, Zhang et al. reported replicable loci in six genes (*EBF1, EEFSEC, AGTR2, WNT4, ADCY5* and *RAP2C*) associated with gestational duration (12). This study identified 3 genes (*EBF1, EEFSEC* and *AGTR2*) strongly associated with PTB in a European ancestry cohort of 43,568 women. However, we did not detect such 6 genes from the reported PTB markers and only identified them as DEGs from few published datasets of PTB. This could be due to the heterogeneous sources of samples used in different PTB studies or be explained in part by the genetic complexity of incomplete gestation induced PTB complications. Here, we collected almost 2,000 PTB-related genes from previous PTB studies. The differences in study design, source and subtype of samples, and statistical methods would be important factors that could account for the diversity of PTB variants and genes among the various studies. Considering the possible association of PTB with other traits, we compared differential gene programs on multiple phenotypes between mother and infant reported in previous studies, and demonstrated a high risk of *CRHBP, ICAM1, DNMT1, CNTLN, PLAGL1, DKK1* and *EGR2* with PTB among our sample cohorts of Chinese or Eastern Asian ancestry. To support this finding, we reconstructed co-expressive networks linking the PTB-related genes in GWAS with the reported PTB markers. Indeed, the correlated PTB genes are mainly involved in immune & inflammation related processes and signaling pathways, as well as coagulation factors. Thus, these findings have biological implications for dissecting genetic associations of gestational factors with disease or traits in different human ancestries.

Pregnancy is associated with an increased risk of developing breast cancer (66). Although several reports indirectly described relationship between breast cancer and PTB (67), evidence is still lacking. To determine whether a correlation exists between PTB women and breast cancer pathogenesis, we compared differential expression of the PTB related genes in PTB samples and three subtypes of hormone receptor-related breast cancer samples. Although several DEGs were found in both PTB and cancer subtypes, their expression patterns are not consistent. It perhaps is because of different subtype of samples derived from heterogeneous populations, as well as different phenotypes involved in the analyses. Nevertheless, our data provides additional evidence that PTB might be related with breast cancer hormone-related subtypes. A graphic overview of our results is summarized in Fig. 6c. We proposed interplays of gestational age with PTB, fetal disease and breast cancer. The representative PTB genes *CRHBP, ICAM1, THBS4, DNMT1, CNTLN, PLAGL1, DKK1* and *EGR2* are likely associated with these phenotypes by targeting immune and inflammatory response, coagulation, and cell adhesion.

## Conclusions

This study benchmarked the ability of ulcWGS to be used for genotype imputation. As the first study using a large cohort of human embryo samples with ulcWGS, we demonstrate its power and effectiveness in GWAS. We detected 40 significant SNPs and 11 genomic risk loci that contains independent significant SNPs and are associated with gestational age. From 166 genes mapped to the risk loci. We establish interrelationships between the mapped genes and maternal or infant diseases and provide insights into understanding the genetic associations of gestational ages. Our findings should expand current GWAS related to gestational duration and preterm trait by including Chinese and East Asian samples and would therefore be helpful to future research.

## Data Availability

The datasets supporting the conclusions of this article are included within the article and its additional files. The significant SNPs and candidate SNPs are listed in Additional file 1: Tables S3 and S4, respectively. The processed or raw counts of mRNA expression datasets shown in Additional file 1: Table S8 was downloaded from Gene Expression Omnibus (GEO)/National Center for Biotechnology Information (NCBI).

## Supplementary Information

**Additional file 1: Table S1.** Genotype imputation performance at different ultra-low coverages and sample sizes. **Table S2**. WGA methods and sequencing platforms used in the PGT experiments. **Table S3**. The summary of 40 significant SNPs satisfying Bonferroni-corrected significant levels of 4.526e-8 from 1,744 embryo PGT samples. **Table S4**. The list of candidate SNPs. **Table S5**. Nonsynonymous SNPs that are independent significant SNPs or in LD (r2>0.6) with one of the independent significant SNPs. **Table S6**. The reported genomic risk loci from GWAS catalog that were detected in our dataset. **Table S7**. A list of 166 mapped genes by positional, eQTL, and chromatin interaction mappings. **Table S8**. Data sources of genome-wide mRNA expression in Preterm birth, infant disease and breast cancer. **Table S9**. A list of the 166 mapped genes that were also identified by differentially expressed genes (DEGs) derived from analyzing the genome-wide gene expression datasets listed.

**Additional file 2: Supplemental Methods**

**Additional file 3: Figure S1.** Sequencing coverage statistics of the 17,844 embryos. **Figure S2.** Visualizing the linear regression model for genotype accuracy prediction. **Figure S3.** Quality control of SNP calling. Figure S4. Figure S4. Principal component analysis of all 17,844 embryo samples. **Figure S5.** Distribution of gestational age in the 1,744 born embryo samples. **Figure S6.** Quantile-Quantile plot of the 1,107,198 studied SNPs. **Figure S7.** Pie charts of th**e** candidate SNPs. **Figure S8.** Zoom in locus plots of the 11 genomic risk loci. **Figure S9.** Circos plots of chromatin interactions and eQTL mapping. **Figure S10.** Gene set analysis of the 166 mapped genes. **Figure 11.** Co-expression network of PTB-related genes in maternal and infant subtypes.

## Abbreviations

WGS: whole genome sequencing
NGS: next-generation sequencing
GWAS: genome-wide association study
bp: base pair
SNP: single nucleotide polymorphism
PTB: preterm birth
PGT: preimplantation genetic testing
MAF: minor allele frequency
LD: linkage disequilibrium
HWE: Hardy-Weinberg equilibrium
WGA: whole genome amplification
PCA: principal component analysis
eQTL: expression quantitative trait locus
FDR: false discovery rate
GO: gene ontology
BPD: bronchopulmonary dysplasia
ER: Estrogen receptor
PR: Progesterone receptor
HER2: human epidermal growth factor receptor 2
TNBC: Triple-negative breast cancer
DEG: differentially expressed gene
AF: allele frequency
GIAB: Genome in a Bottle
BWA: Burrows-Wheeler Aligner
GEO: Gene Expression Omnibus
NCBI: National Center for Biotechnology Information

## Declarations

### Ethics approval and consent to participate

This study has been reviewed and approved by the Institutional Review Board (IRB) of China International Trust Investment Corporation - Xianngya (IRB Reference No. LL-SC-2020-004). The study used data for clinical PGT samples and their complete follow-up records collected at the Reproductive & Genetic Hospital of CITIC-XIANGYA between 2013 and 2019 for retrospective analysis. All patients provided written informed consent for PGT and medical research. Following the regulations of the Human Genetic Resources Administration of China, all genetic materials involved in this study have been reviewed and approved by the Ministry of Science and Technology of the Peoplés Republic of China (Approval No. [2022] GH1831).

### Consent for publication

Not applicable

## Funding

The study was supported by the Early Career Schema (27204518) of the Hong Kong Research Grants Council for RL, partially by General Research Funding of Hong Kong Research Grants Council (17113721) for RL and (17117918) for BY, by the University Grants Committees Fund from the University of Hong Kong for RL, partially by National Key Research and Developmental Program of China (2018YFC1004900) for YT, by the Innovation and Technology Fund (ITF/331/17FP) of Innovation and Technology Commission of the Hong Kong SAR government for TL.

### Authors’ contributions

SL, RL and BY developed the computational pipeline and the algorithms. RL, GL, YW, TKL, and TL designed the experiments. PX, YG, FG and YT contributed to the clinical data collection. SL, BY, JL, YG and FG performed data processing and analysis. RL, GL, SL, BY, PX and YW participated in drafting the manuscript. All authors read and approved the final manuscript.

## Acknowledgements

Not applicable

