## Supplementary Methods for "Ultra-low coverage genome-wide association study – insights into gestational age using 17,844 embryo samples with preimplantation genetic testing"

### 1. **SNP Calling**

#### 1.1. **Maximum log-likelihood estimation of allele frequency**

We improved the method of allele frequency calculation of a recent study by using log-likelihood to handle ultra-low coverage and very large sample size without suffering from precision loss under the current computer architecture (1). First, for one chromosome site, we randomly selected one read for each of the samples and used *samtools mpileup -Q20* to pile up the bases with a base quality score larger than 20. The log-likelihood of the allele frequency of a candidate variant site is defined as:

$$p=(p_{A},p_{T},p_{C},p_{G})$$

(1)

$$\ln\left( L\left( p \right) \right)=\sum_{i=1}^{N} ln(P(D_{i}|p))=\sum_{i=1}^{N} ln(\sum_{b\in\{A,T,C,G\}} p_{b}p(D_{i}|b))$$

(2)

$$p\left( D_{i} | b \right)=\left\{ \begin{aligned} 1-\varepsilon_{i},\mathrm{if} D_{i}=b \\ \frac{\varepsilon_{i}}{3},\mathrm{else} \end{aligned} \right.$$

(3)

where *N* is the population size, *p* denotes the allele frequencies at the site and $p_{b}$ is the allele frequency of allele *b* and $b\in(A,T,C,G)$. $D_{i}$ is the sampled base of the *i*^th^ sample at the site. $\varepsilon_{i}$ is the sequencing error rate converted from the PHRED-scale quality of the selected read with $D_{i}$ at the site.

***EM optimization***

The allele frequency of a variant candidate site is estimated by maximizing ln(L(p)) (**Eq. 2**) through Expectation-Maximization (EM) optimization, which is the same with (1). First, the allele frequency *p* is initialized by:

$$p_{b}=\frac{\sum_{i}^{N} D_{i}=b}{N}$$

(4)

In the expectation step, we calculate the posterior probability $p\left( b | D_{i} \right)$ of the expected allele frequency at the candidate variant site, given by the observed nucleotide bases:

$$p\left( b | D_{i} \right)=\frac{p_{b}p(D_{i}|b)}{\sum_{b^{'}\in\{A,T,C,G\}} p_{b}p(D_{i}|b^{'})}$$

(5)

In the maximization step, the allele frequency is updated by maximizing the likelihoods.

$$p_{b}^{'}=\frac{\sum_{i=1}^{N} p\left( b | D_{i} \right)}{N}$$

(6)

The algorithm will be terminated if the change in ln(L(p)) is less than 0.001 or the iteration number has reached 2000.

#### 1.2. **Log-likelihood ratio test**

The aim of the log-likelihood ratio test is to remove the potential noises (such as very low allele frequencies) resulting from the maximum log-likelihood estimation of allele frequency. Instead of using a downstream way to test the hypothesis, we used the estimated allele frequency obtained from maximum log-likelihood estimation as the alternate hypothesis all the time.

The estimated allele frequency was considered as the alternate hypothesis, which was formulated as:

$$p^{\mathrm{alt}}=(p_{A}^{\mathrm{alt}},p_{C}^{\mathrm{alt}},p_{G}^{\mathrm{alt}},p_{T}^{\mathrm{alt}})$$

(7)

$$V_{init}=\sum_{b\in(A,T,C,G)} x_{b},x_{b}=\left\{ \begin{aligned} 1, p_{b}^{alt}>0 \\ 0,\mathrm{else} \end{aligned} \right.$$

(8)

where $p^{\mathrm{alt}}$ denotes the allele frequency *p* derived from maximum log-likelihood estimation, which is used as alternate hypothesis in the log-likelihood ratio test. $p_{b}^{\mathrm{alt}}$ is the estimated allele frequency of base *b,* $b\in(A,T,C,G)$*.* $x_{b}$is encoded as 0 or 1, meaning whether the allele *b* is regarded as a candidate variant. And $V_{init}$ denotes the initial allelic type of the candidate site (1 for mono-allelic site, 2 for bi-allelic site, 3 for tri-allelic site, 4 for tetra-allelic site). The candidate sites with $V_{init}$ > 1 will be used for log-likelihood ratio test. Then, the alternate hypothesis will test against the null hypothesis in a prioritized order using the following strategies.

***1 VS X***

First of all, we generate all possible mono-allelic models as the null hypothesis. The *i*^th^ null hypothesis can be denoted as:

$$p^{{null}_{i}^{'}}=\left( p_{b}^{{null}_{i}^{'}},0,0,0 \right), if p_{b}^{alt}>0$$

(9)

$$p_{b}^{{null}_{i}^{'}}=p_{b}^{alt}$$

(10)

$p^{{null}_{i}^{'}}$ is normalized to 1 by:

$$p^{{null}_{i}}=\frac{p_{b}^{{null}_{i}^{'}}}{\sum_{b\epsilon\{A,T,C,G\}} p_{b}^{{null}_{i}^{'}}}$$

(11)

where $p^{{null}_{i}}$ means the allele frequency for the *i*^th^ null hypothesis. In our case, the number of null hypothesis equals $V_{init}$.

Then, the estimator of the log-likelihood ratio test is shown as:

$$\mathrm{estimator}=-2[ln(L\left( p^{alt} \right)-ln(L(p^{{null}_{i}})))$$

(12)

where $ln(L\left( p \right))$ is formulated in **Eq. 2**.

We let the estimator follow a chi-square distribution. The degree of freedom is determined by how many different parameters are between the null and alternate hypothesis. Thus, in this case, the degree of freedom is $V_{init}$.

This test would be applied to all possible null hypotheses. Then, we select the $p^{{null}_{i}}$ that produces the maximum p value. If this p value is larger than the threshold of 1e-6, the process is terminated here and the candidate site is regarded as a mono-allelic site. If the p value is smaller than the threshold, and $V_{init}=2$, then the process is also terminated here and the candidate site is regarded as a bi-allelic site. Otherwise, it is considered there should be at least 2 variants and the process will proceed to the next procedure.

***2 VS X***

If the maximum p value of 1 vs X is smaller than the threshold and $V_{init}>2$, we will continue to generate all possible bi-allelic models as the null hypothesis and test them against the alternate hypothesis $p_{b}^{alt}$.

We generated the null hypothesis of the allele frequency based on the following rules:

$$p^{{null}_{i}^{'}}=\left( p_{b1}^{{null}_{i}^{'}},p_{b2}^{{null}_{i}^{'}},0,0 \right),\mathrm{if} p_{b1}^{alt}>0 \mathrm{and} p_{b2}^{alt}>0$$

(13)

$$b_{1}, b_{2}\in\{A,T,C,G\}$$

(14)

$$p_{b1}^{{null}_{i}^{'}}=p_{b1}^{alt},p_{b2}^{{null}_{i}^{'}}=p_{b2}^{alt}$$

(15)

$p^{{null}_{i}^{'}}$ is normalized to 1 using **Eq. 11**. Then, log-likelihood ratio test is applied to test all possible bi-allelic models against the alternate hypothesis using **Eq. 12**.

If the maximum p value is larger than the threshold of 1e-6, the process is terminated here, and the candidate site is regarded as a bi-allelic site. If this p value is smaller than the threshold, and $V_{init}=3$, the process is also terminated here and the candidate site is regarded as a tri-allelic site. Otherwise, it is considered there should be at least 3 variants and the process will proceed to the next procedure.

***3 VS X***

We generated all possible tri-allelic models based on the following rules:

$$p^{{null}_{i}^{'}}=\left( p_{b1}^{{null}_{i}^{'}},p_{b2}^{{null}_{i}^{'}},p_{b3}^{{null}_{i}^{'}},0 \right),\mathrm{if} p_{b1}^{alt}>0 , p_{b2}^{alt}>0, p_{b3}^{alt}>0$$

(16)

$$b_{1}, b_{2}, b_{3}\in\{A,T,C,G\}$$

(17)

$$p_{b1}^{{null}_{i}^{'}}=p_{b1}^{alt},p_{b2}^{{null}_{i}^{'}}=p_{b2}^{alt},p_{b3}^{{null}_{i}^{'}}=p_{b3}^{alt}$$

(18)

$p^{{null}_{i}^{'}}$is normalized to 1 using **Eq. 11**. Then, log-likelihood ratio test is applied to test all possible tri-allelic models against the alternate hypothesis based on **Eq. 12**.

If the maximum p value is larger than the threshold of 1e-6, the process is terminated here, and the candidate site is regarded as a tri-allelic site. Otherwise, it will be considered as the tetra-allelic site, indicated by the alternate hypothesis.

### 2. **Principal Component Analysis (PCA)**

In our study we used Principal Component Analysis in two analyses. 1) We identified potential biases using all 17,844 embryo datasets at the dbSNP known SNPs with MAF≥0.05 (Additional file 3: Figure S4) and concluded that the two most significant principal components represent different whole genome amplification kits, and sequencing platforms. 2) So, to remove the biases, we performed PCA on imputed and filtered SNPs in the 1,744 born embryos with gestational age, and used the eight most significant principle components as covariates to be considered in subsequent GWAS analyses.

The genotype probabilities calculated by STITCH (2) were used for calculating the covariance matrix as:

$$h_{m}^{i}=\sum_{k=0,1,2} \frac{1}{2}kP_{k}$$

(19)

$$C_{i,j}=\frac{1}{M}\sum_{m=1}^{M} \frac{(h_{m}^{i}-f_{m})(h_{m}^{j}-f_{m})}{f_{m}(1-f_{m})}$$

(20)

where $P_{k}$ denotes the imputed genotype probability for genotype *k*, with *k*=0 means genotype 0/0, *k*=1 means genotype 0/1 or 1/0, and *k*=2 means genotype 1/1. $f_{m}$ is the minor allele frequency of the *m*^th^ SNP and *M* is the total number of SNPs. $C_{i,j}$ is the covariance matrix.

With over ten thousand samples and a hundred million SNPs, it was infeasible to get the PCA done in a reasonable time on CPU. Instead, we calculated the covariance matrix using CuPy (3) on GPU. The decomposition of the covariance matrix was calculated by the SciPy package (4).
