## Supplementary Figures for "Ultra-low coverage genome-wide association study – insights into gestational age using 17,844 embryo samples with preimplantation genetic testing"

a

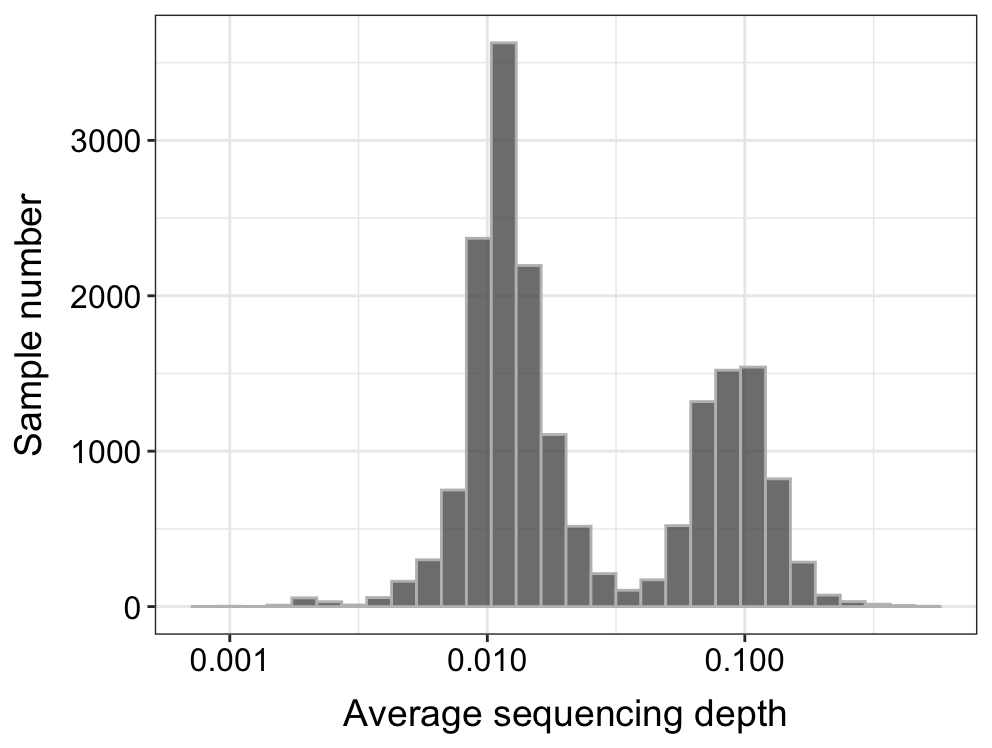

b

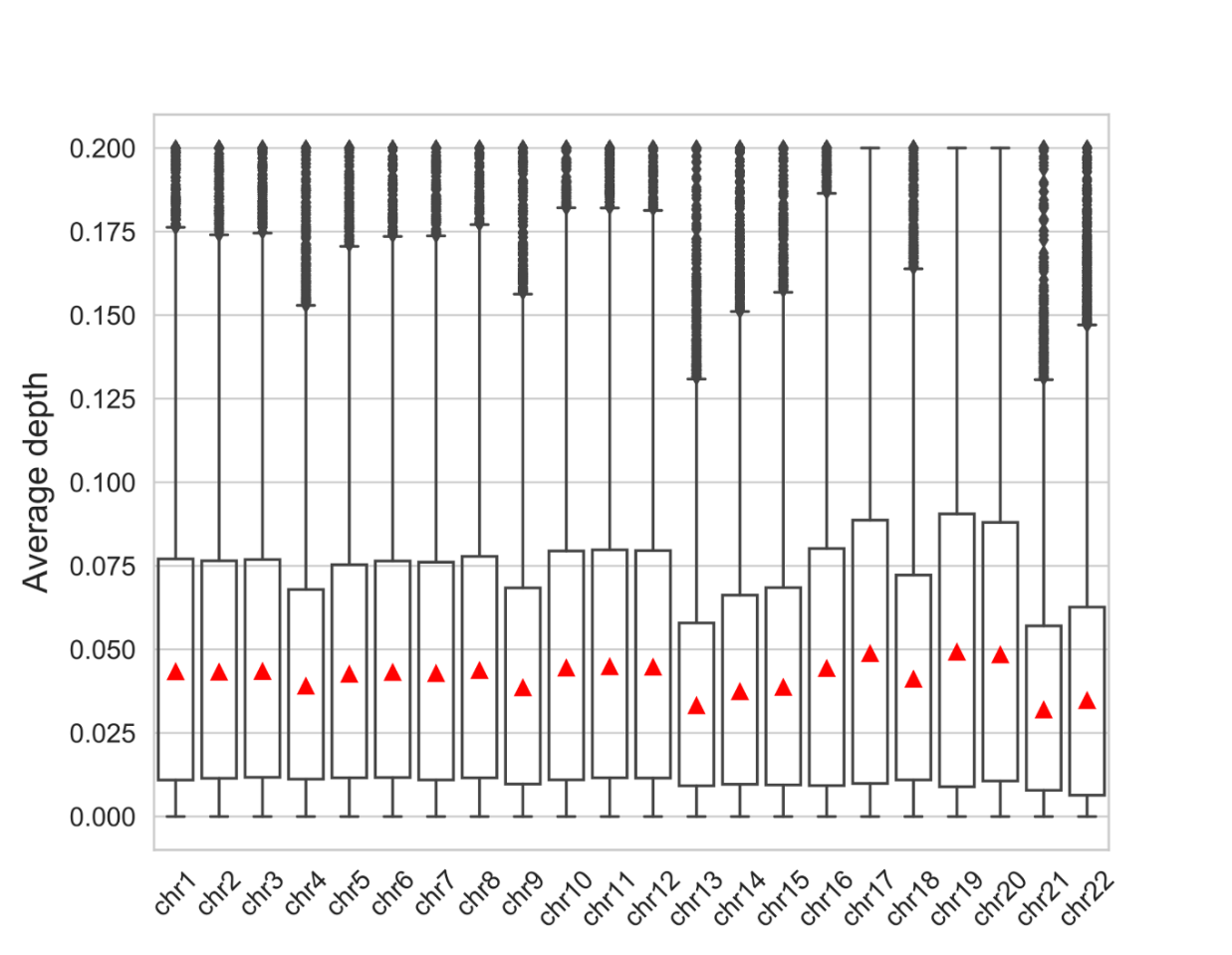

**Figure S1.** **Sequencing coverage statistics of the 17,844 embryos.** a, the sequencing coverage distribution of samples (of 17 with sequencing coverage below 0.001 are not shown). b, the sequencing coverage of each autosome. The red triangle denotes average sequencing coverage.

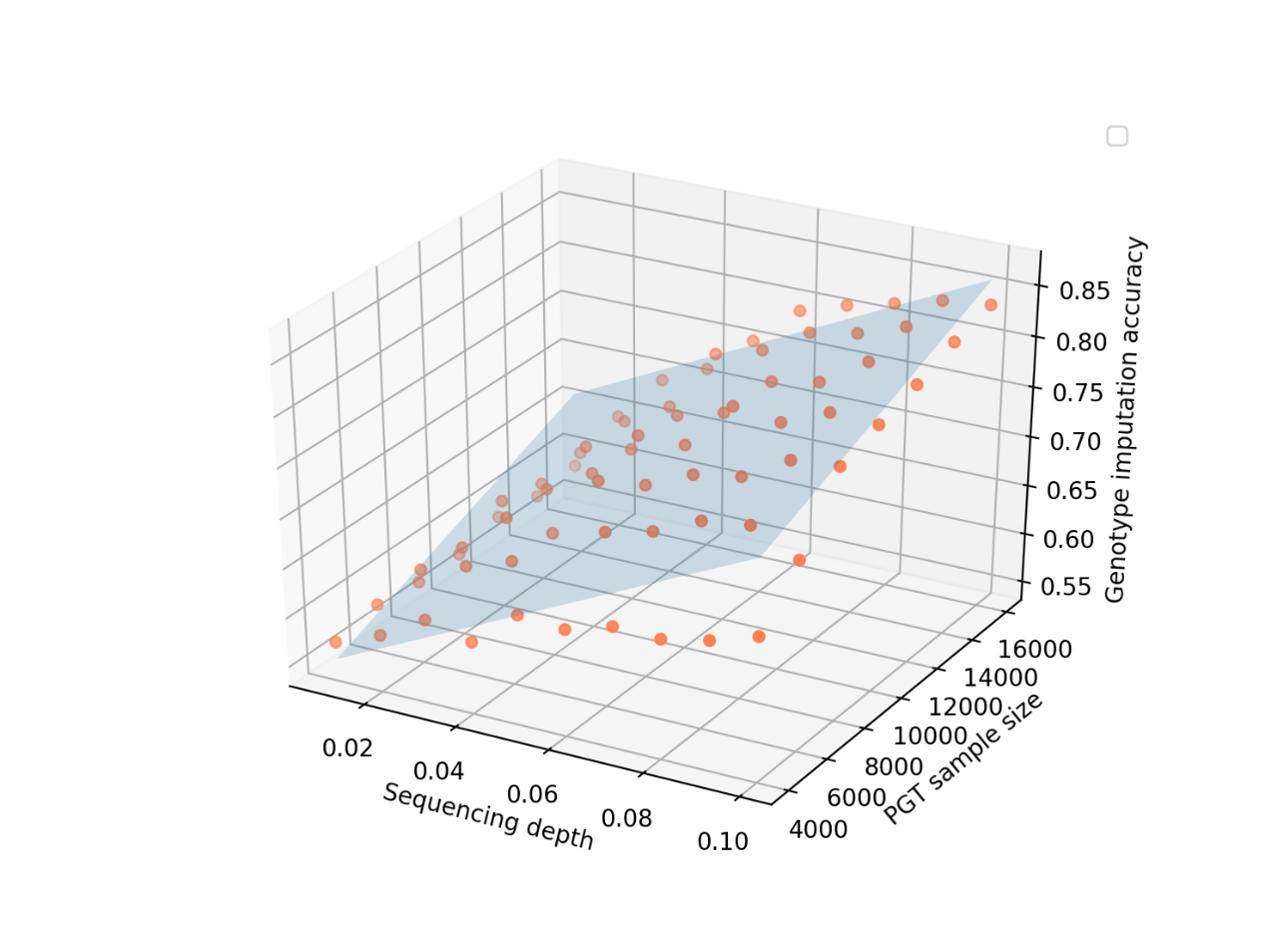

**Figure S2.** **Visualizing the linear regression model for genotype accuracy prediction.** The linear regression model**:** $acc=2.227*c+8.937e^{-6}*s+0.494 (c\geq0.01,s\geq4000)$, where *acc* is the expected genotype accuracy, *c* denotes the sequencing coverage, and *s* denotes sample size.

a

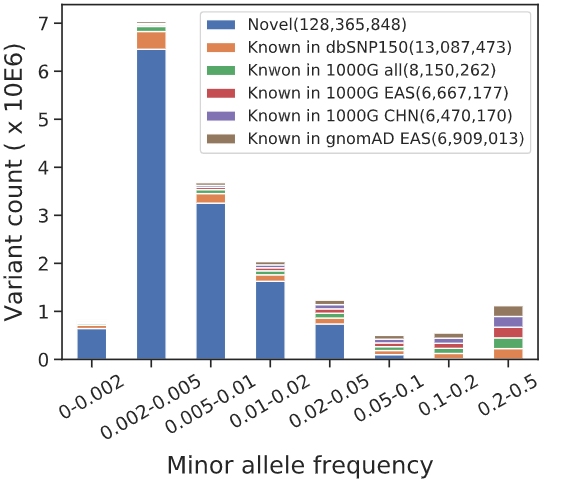

b

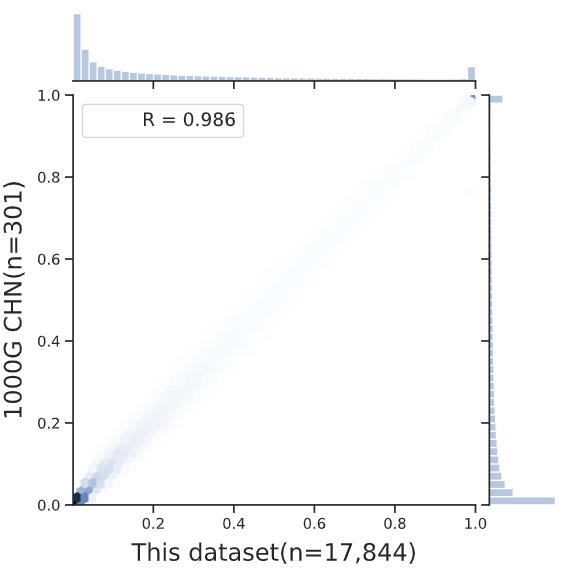

**Figure S3.** **Quality control of SNP calling.** a, the allele frequency spectrum of bi-allelic SNPs in 17,844 embryo samples**.** b, Pearson correlation coefficient of non-reference allele frequencies between 1000G CHN and the 17,844 embryo dataset. Known SNPs with $\mathrm{MAF}\geq0.01$ were included.

a

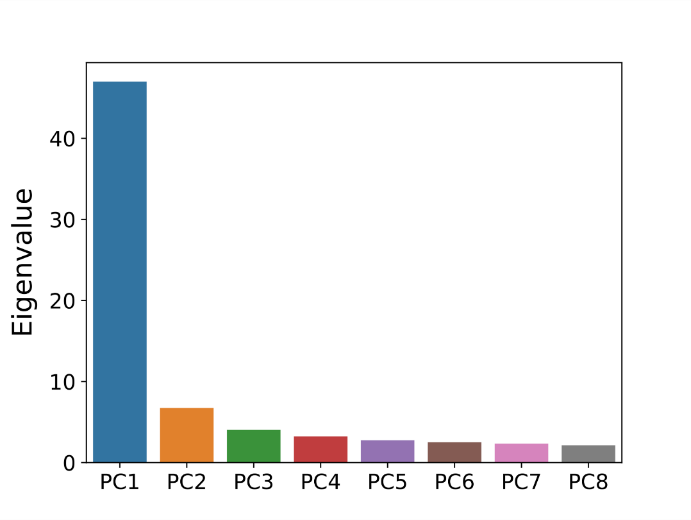

b

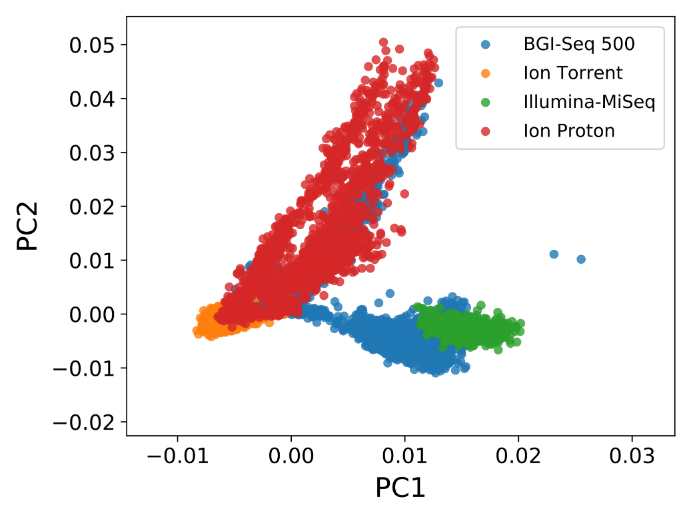

c

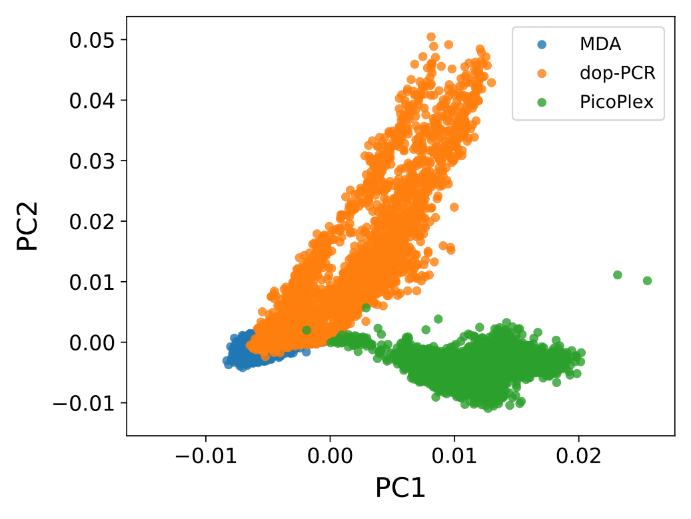

**Figure S4. Principal component analysis of all 17,844 embryo samples.** Imputed genotypes of SNPs with MAF ≥ 0.05 were used as the input. a, the eigenvalue of the top 8 principal components (PCs). b, the samples are colored by sequencing platforms, each dot is a sample. c, the samples are colored by WGA methods and each dot is a sample.

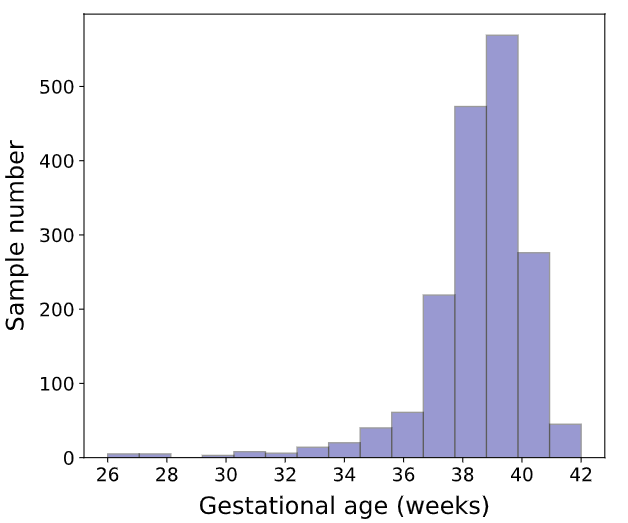

**Figure S5.** **Distribution of gestational age in the 1,744 born embryo samples.**

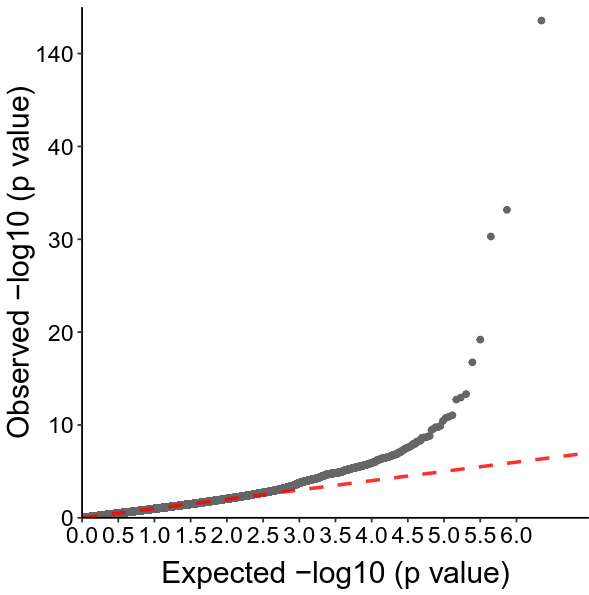

**Figure S6. Quantile-Quantile plot of the 1,107,198 studied SNPs.**

a

Chromatin state

|  | percentage |
| --- | --- |
| TssA | 0.0641399 |
| TssAFlnk | 0.0153061 |
| TxFlnk | 0.00218659 |
| Tx | 0.215743 |
| TxWk | 0.355685 |
| Enh | 0.0495627 |
| ZNF/Rpts | 0.0138484 |
| Het | 0.185131 |
| ReprPC | 0.0116618 |
| ReprPCWk | 0.0750729 |
| Quies | 0.0116618 |

b

RegulomeDB

|  | Percentage |
| --- | --- |
| 1d | 0.000728863 |
| 1f | 0.00145773 |
| 2a | 0.000728863 |
| 2b | 0.0102041 |
| 3a | 0.0182216 |
| 4 | 0.03207 |
| 5 | 0.137755 |
| 6 | 0.280612 |
| 7 | 0.347668 |
| None | 0.170554 |

**Figure S7.** **Pie charts of the candidate SNPs.** a, distribution of the candidate SNPs in the 15 core chromatin states derived from 127 epigenomes. b, RegulomeDB scores of candidate SNPs. RegulomeDB score ranging from 1a to 7, indicates the likelihood of these SNPs to affect regulatory process. A score 1a refers the highest likelihood, and 7 means that there is no evidence that the SNPs function as regulatory elements.

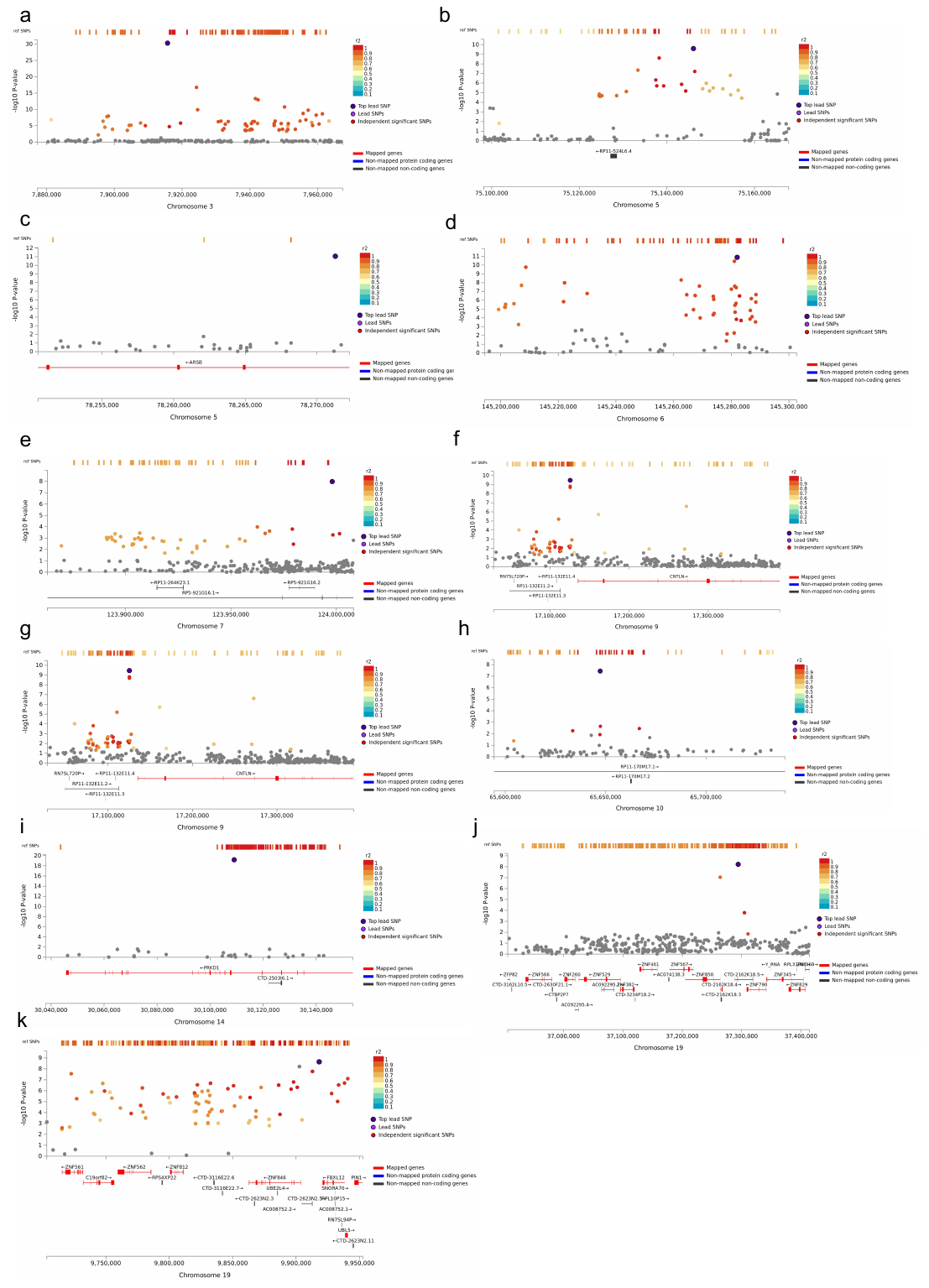

**Figure S8. Zoom in locus plots of the 11 genomic risk loci.**

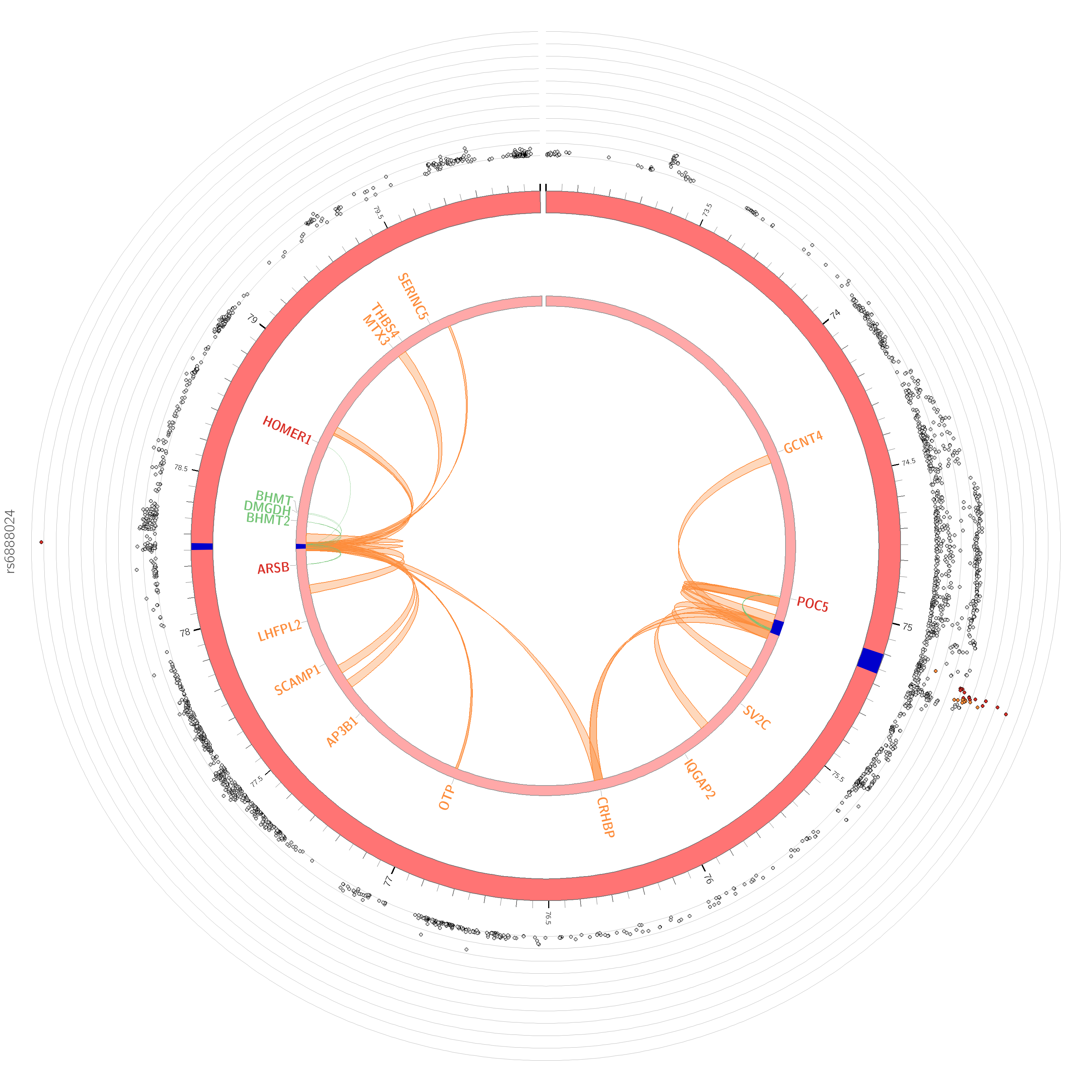

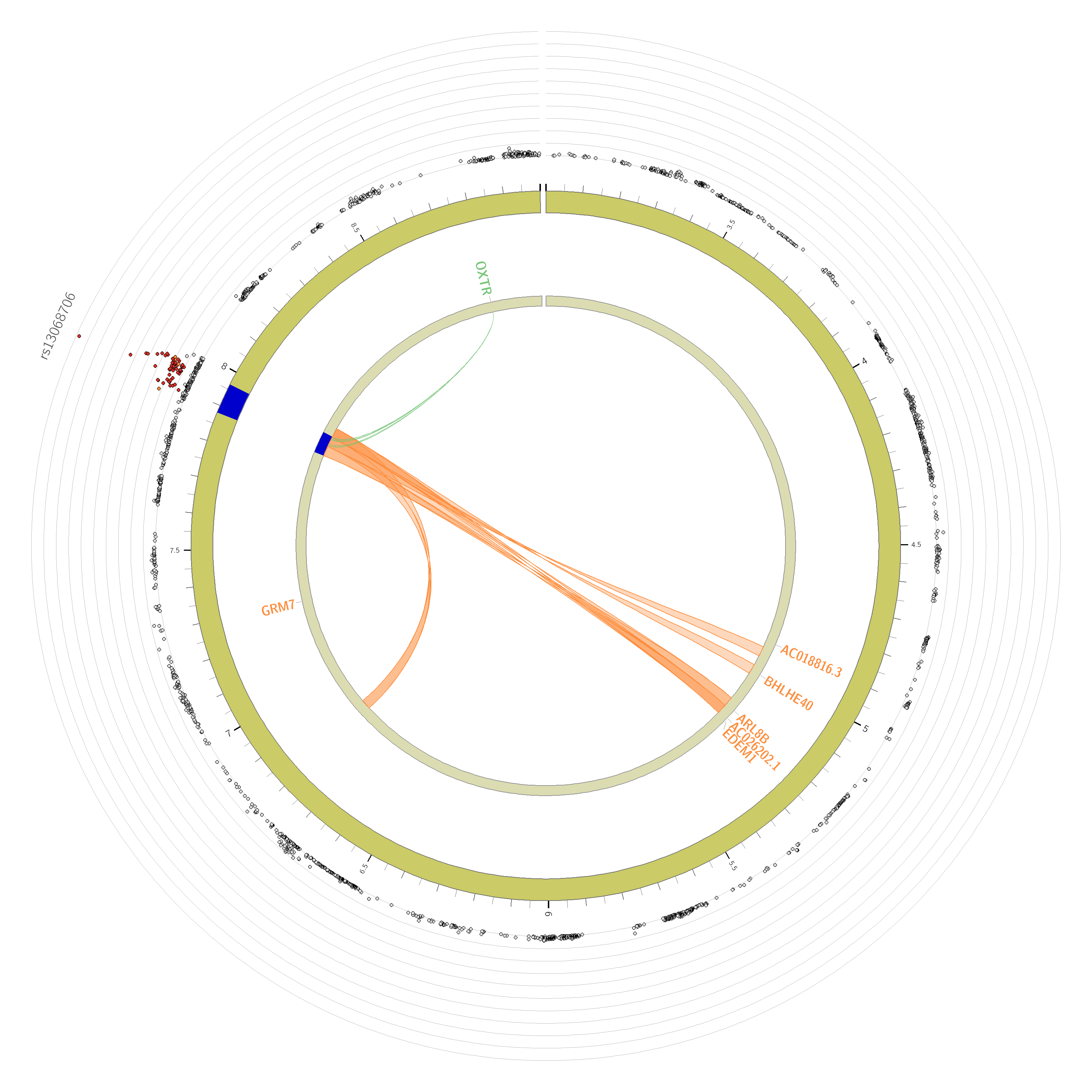

a

b

Chromosome 3

Chromosome 5

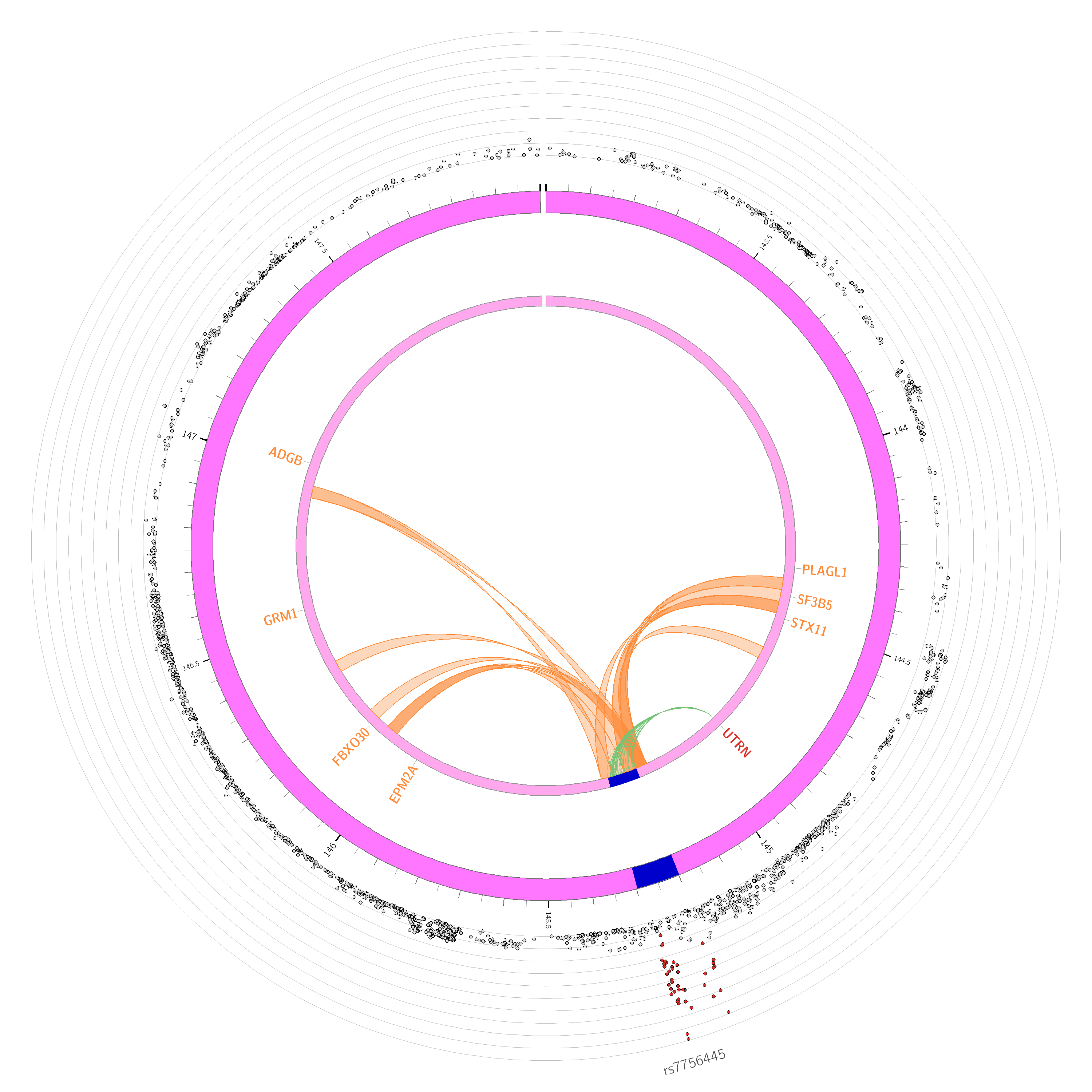

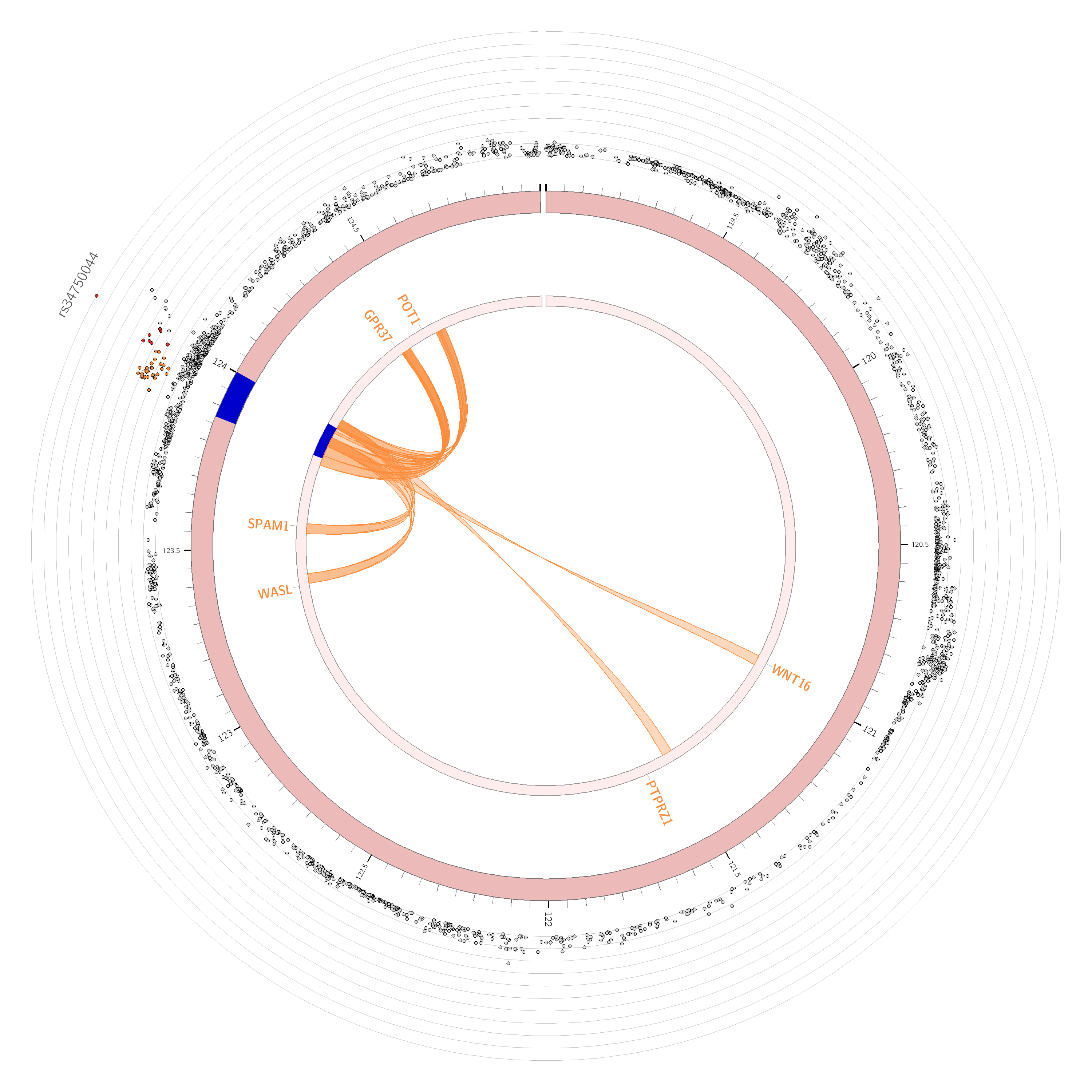

c

d

Chromosome 6

Chromosome 7

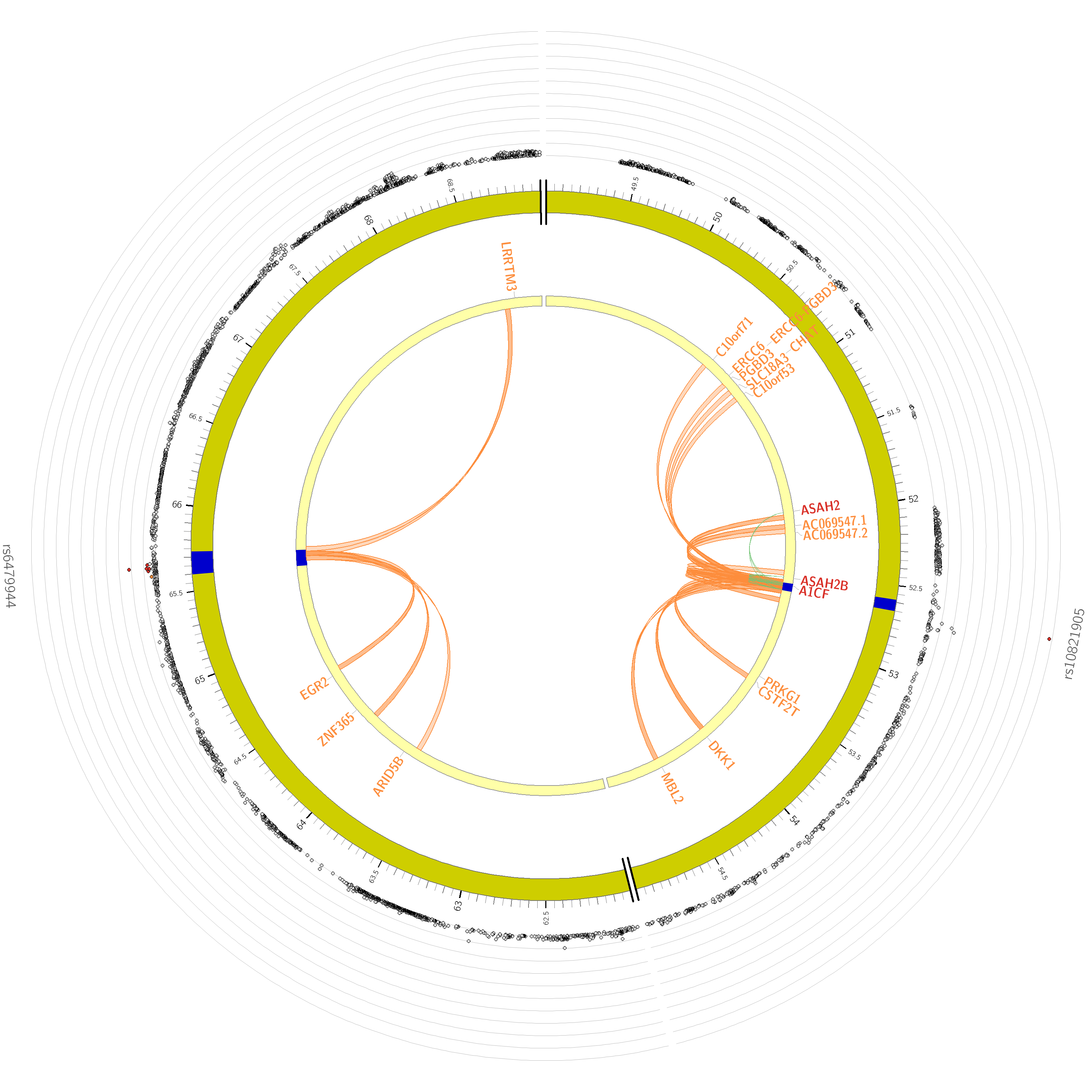

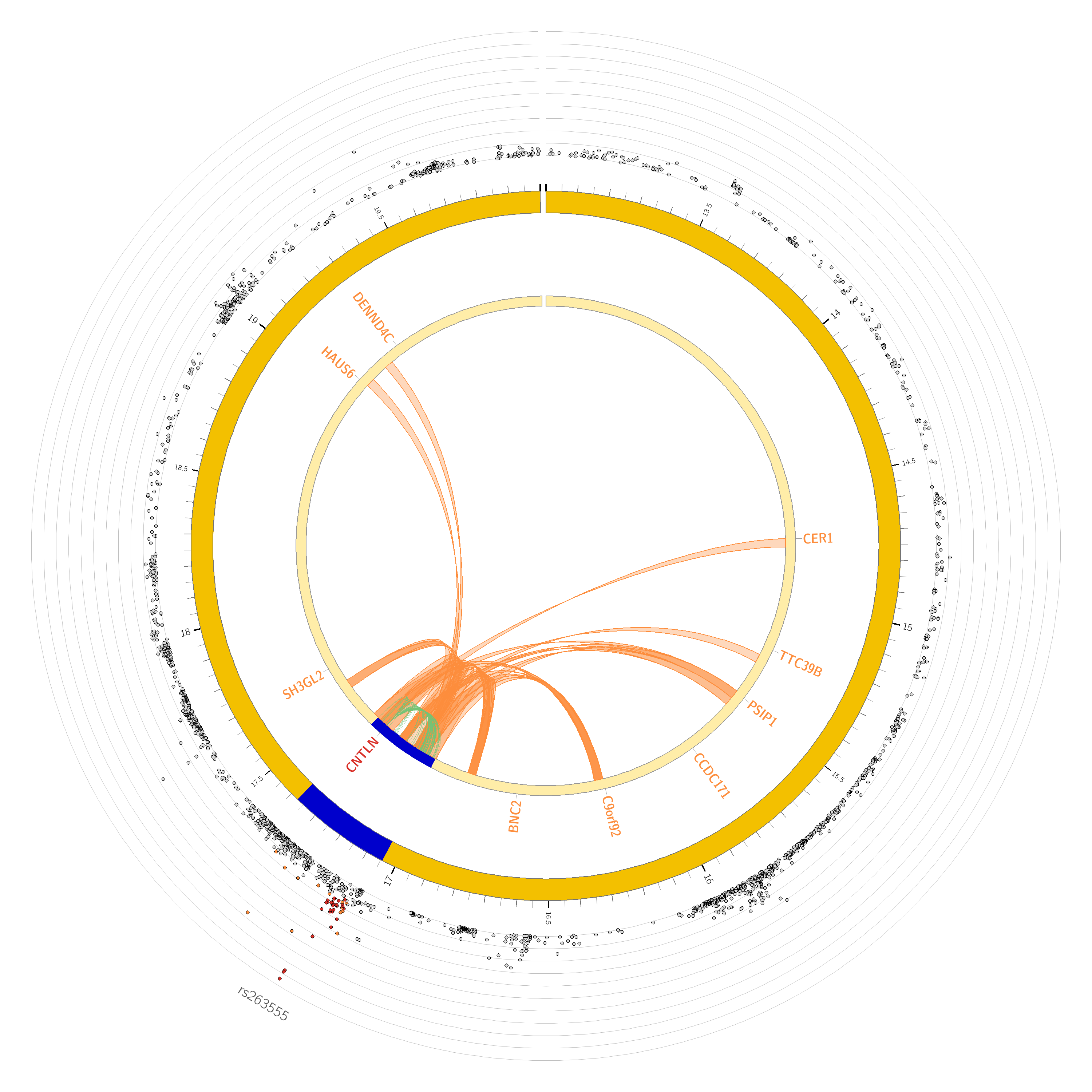

e

f

Chromosome 9

Chromosome 10

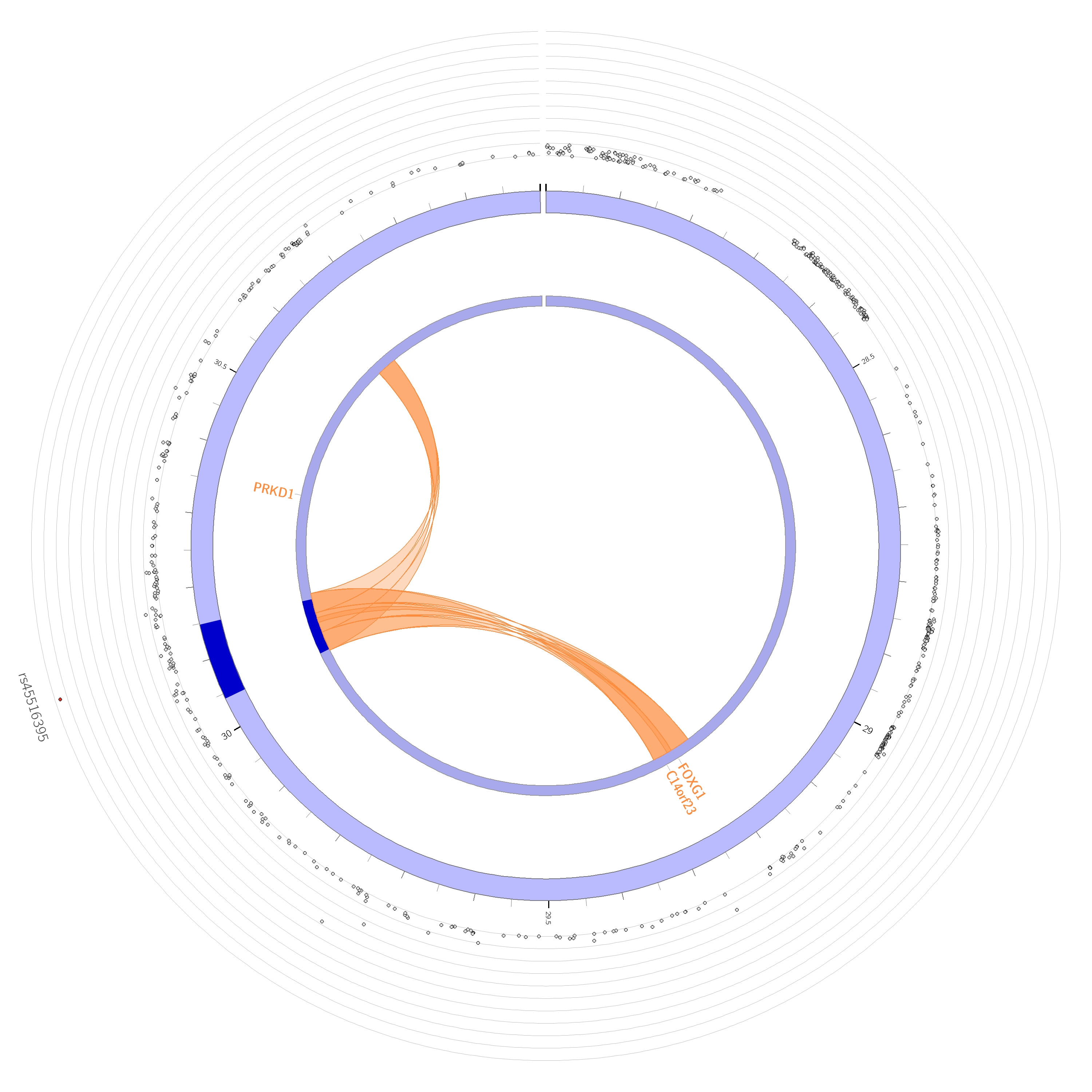

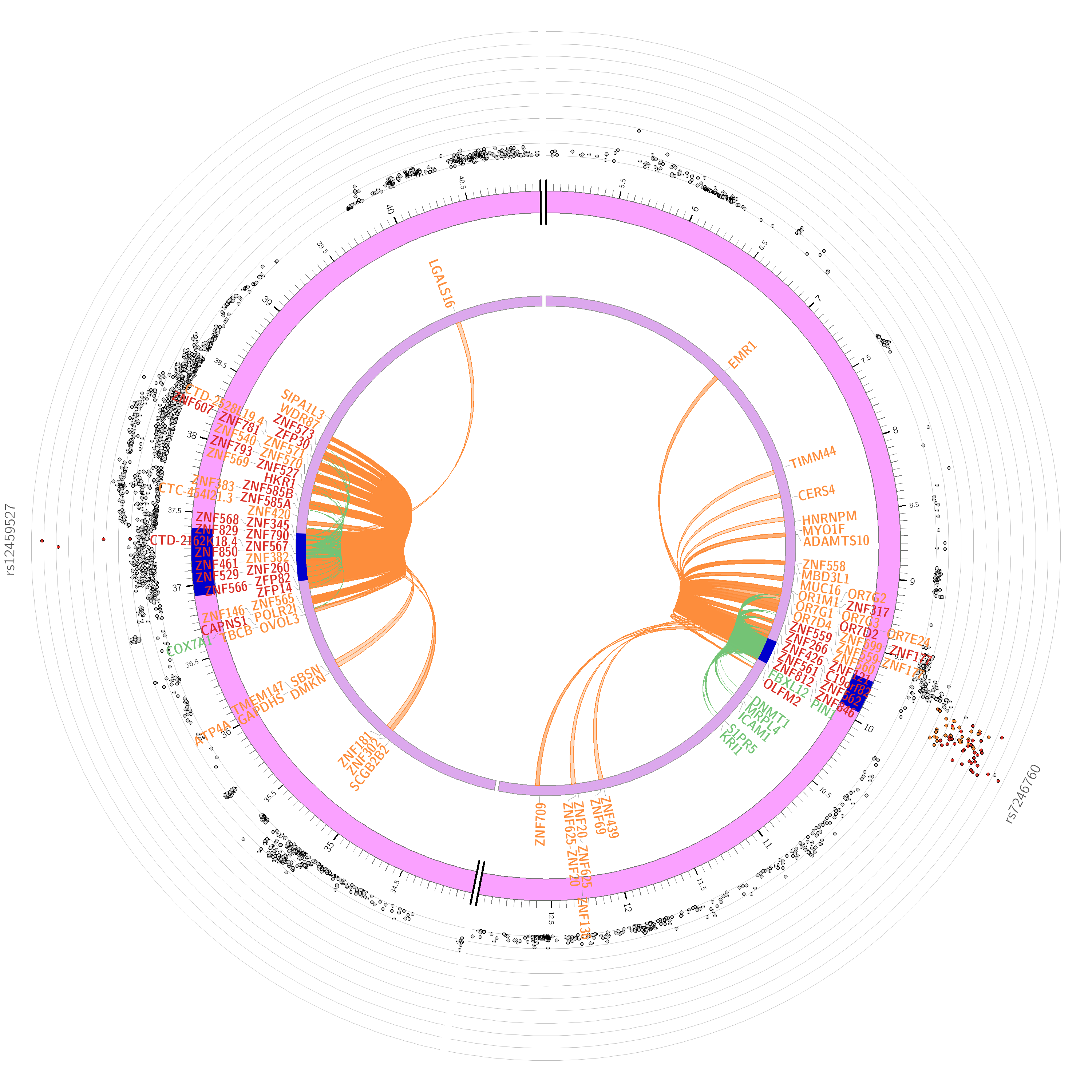

g

h

Chromosome 14

Chromosome 19

**Figure S9.** **Circos plots of chromatin interactions and eQTL mapping.** The first outer ring is the Manhattan plot showing the SNP. The color of SNPs denotes their maximum $r^{2}$ with another independent significant SNP within the genomic risk loci, red for $r^{2}$ > 0.8, orange for $r^{2}$ > 0.4, blue for $r^{2}$ > 0.2 and others are grey. The second outer ring is a chromosome, with blue regions denoting genomic risk loci. The genes mapping methods or strategies are shown in different colors, green by eQTL, orange by chromatin interaction and red by both. The inner ring shows the links with eQTL association (in green) or with chromatin interaction (in orange).

a

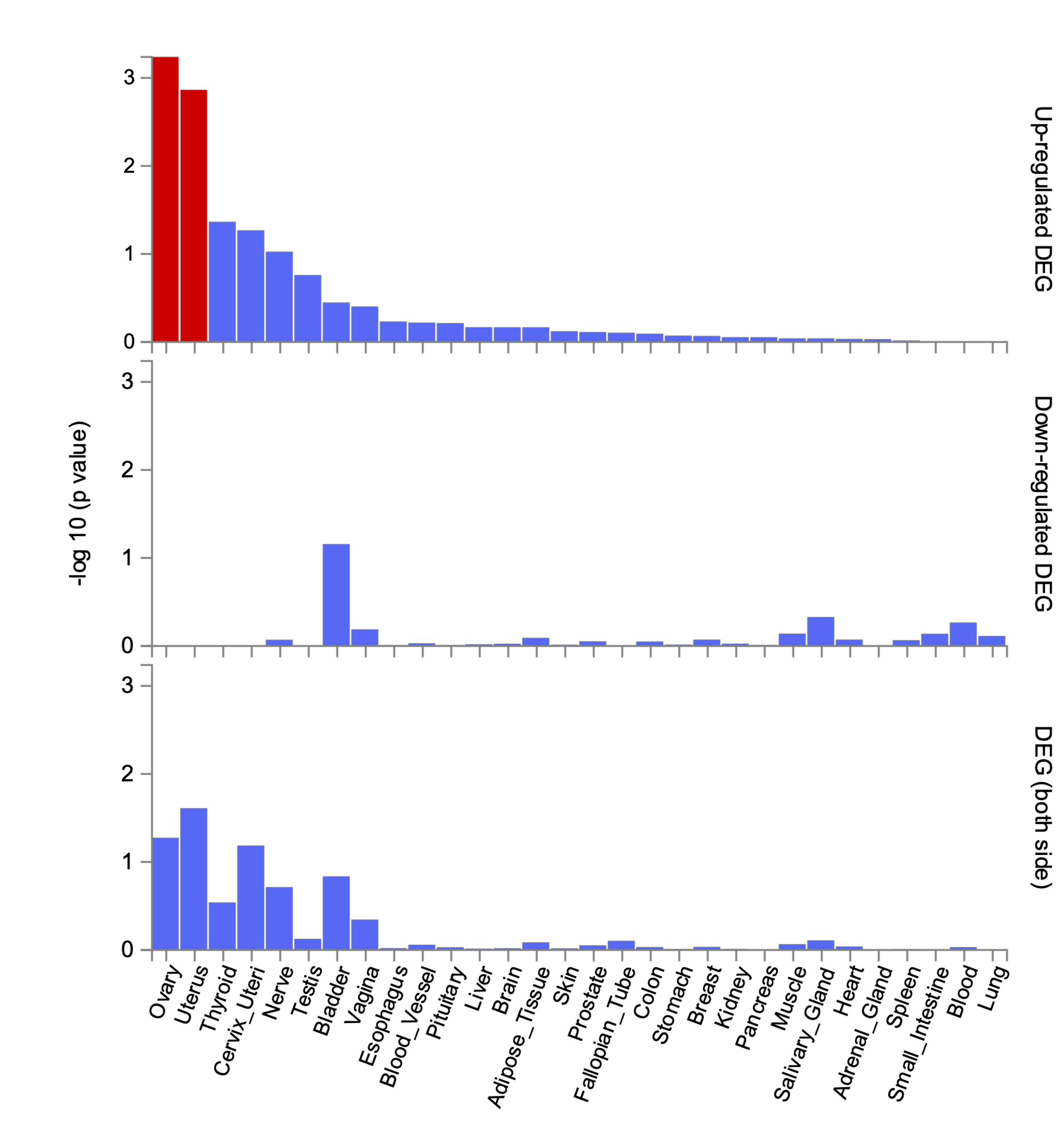

b

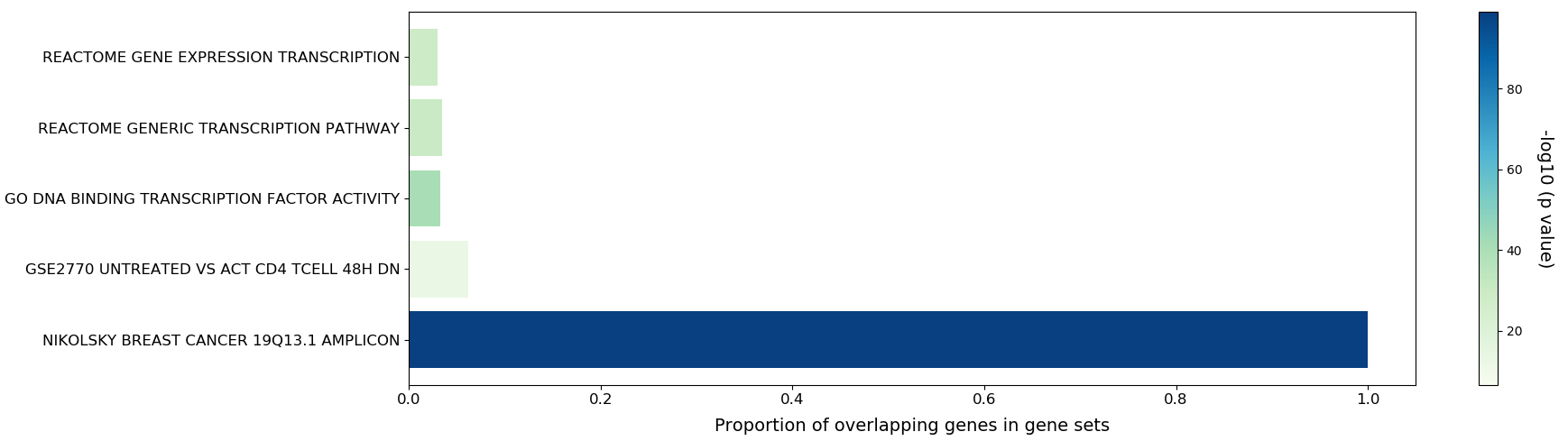

**Figure S10. Gene set analysis of the 166 mapped genes.** a, enrichment of differentially expressed genes sets in GTEx v8 general tissue types. b, the results of gene-set enrichment analysis by using different gene sets from MSigDB (<https://www.gsea-msigdb.org/gsea/msigdb/>) as background.

a
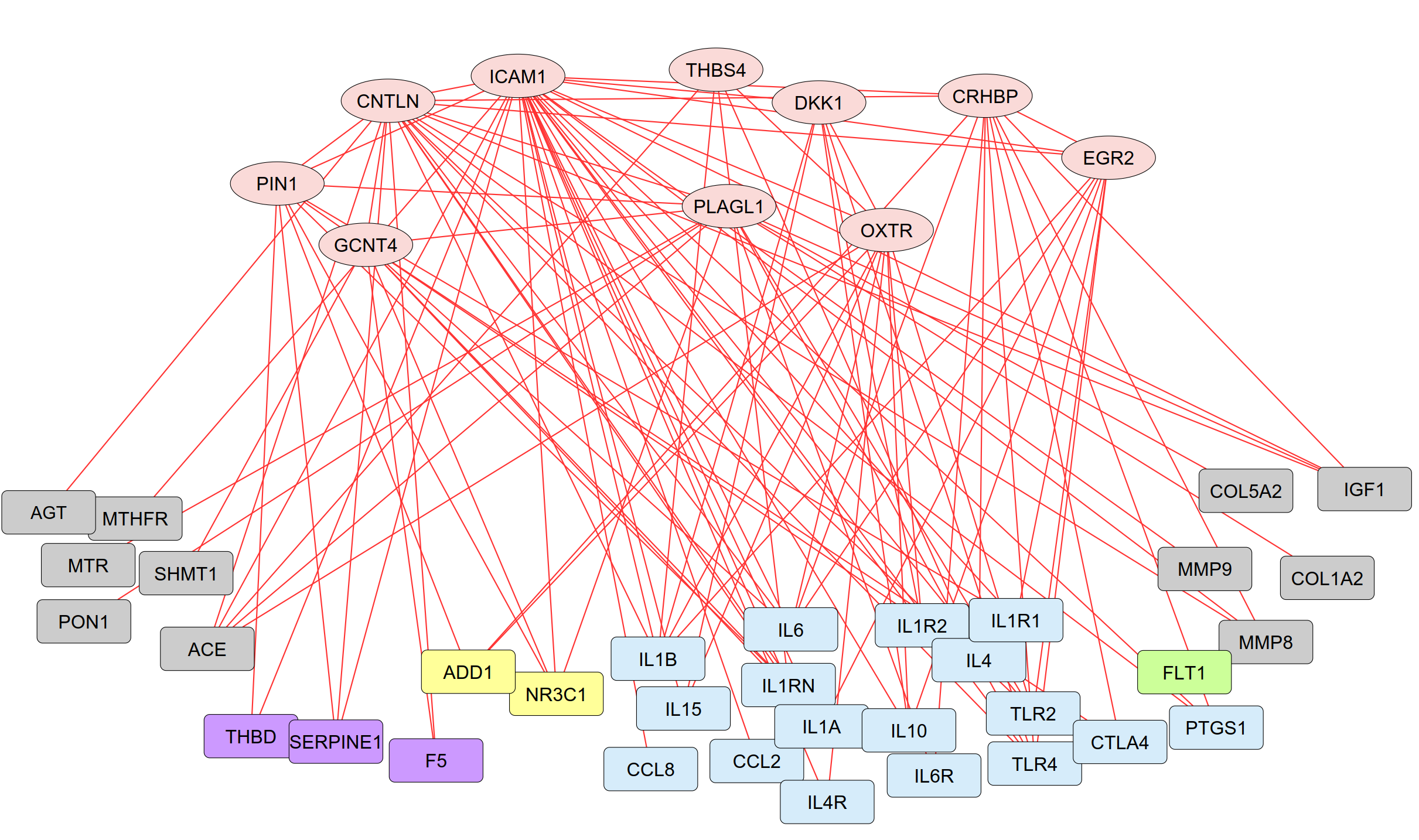

b

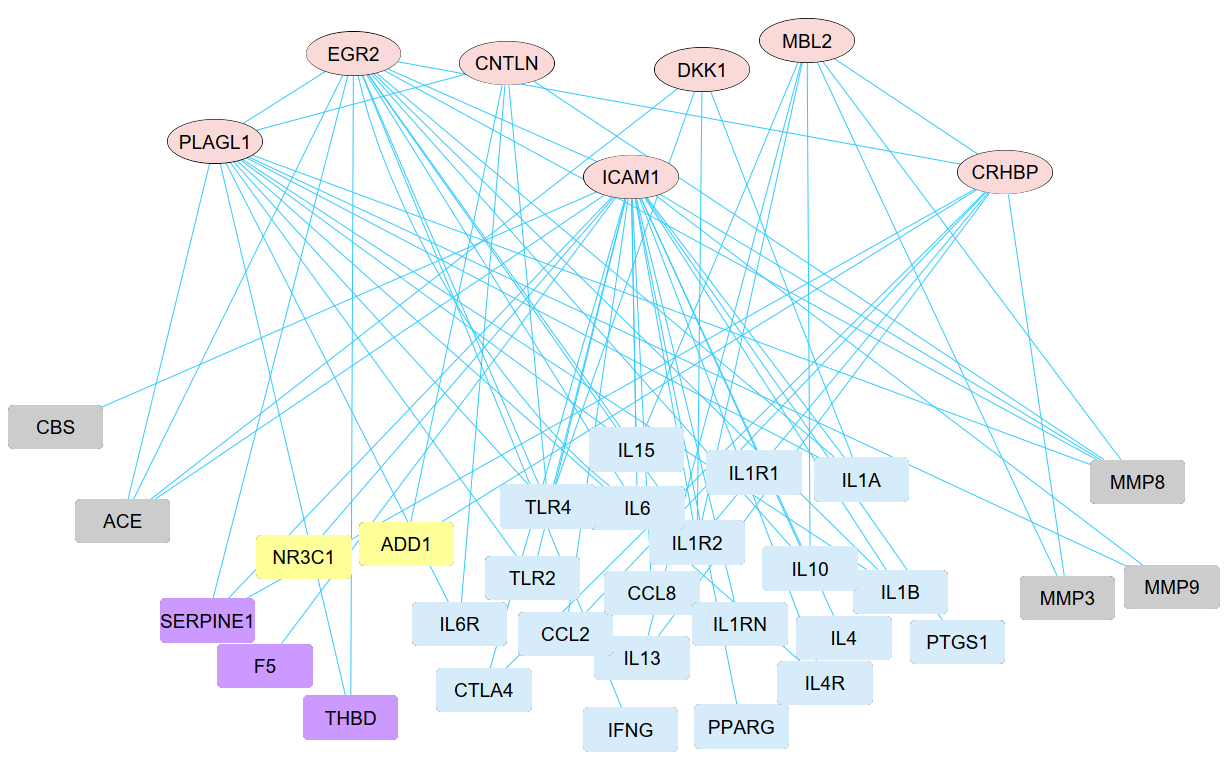

c

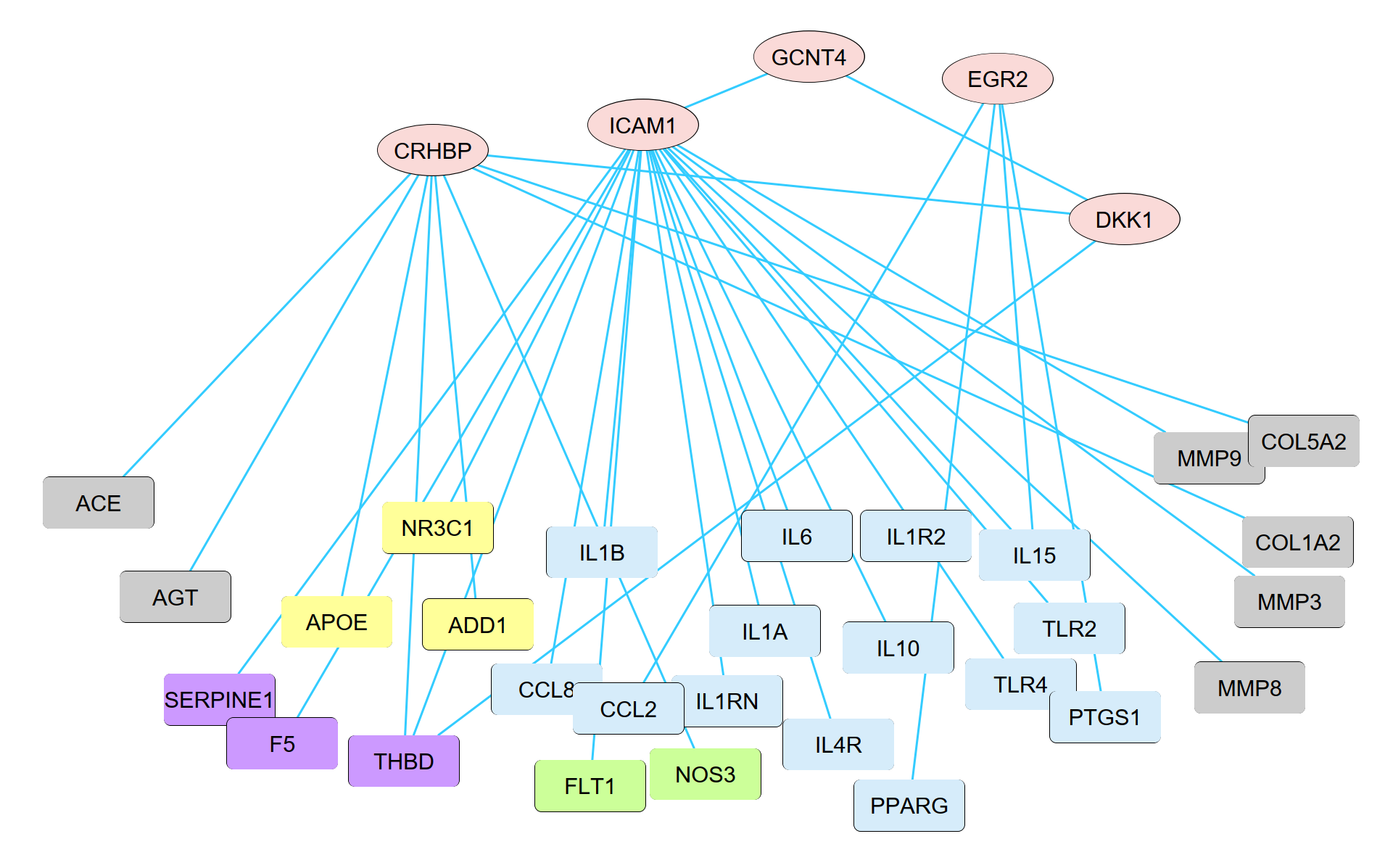

d

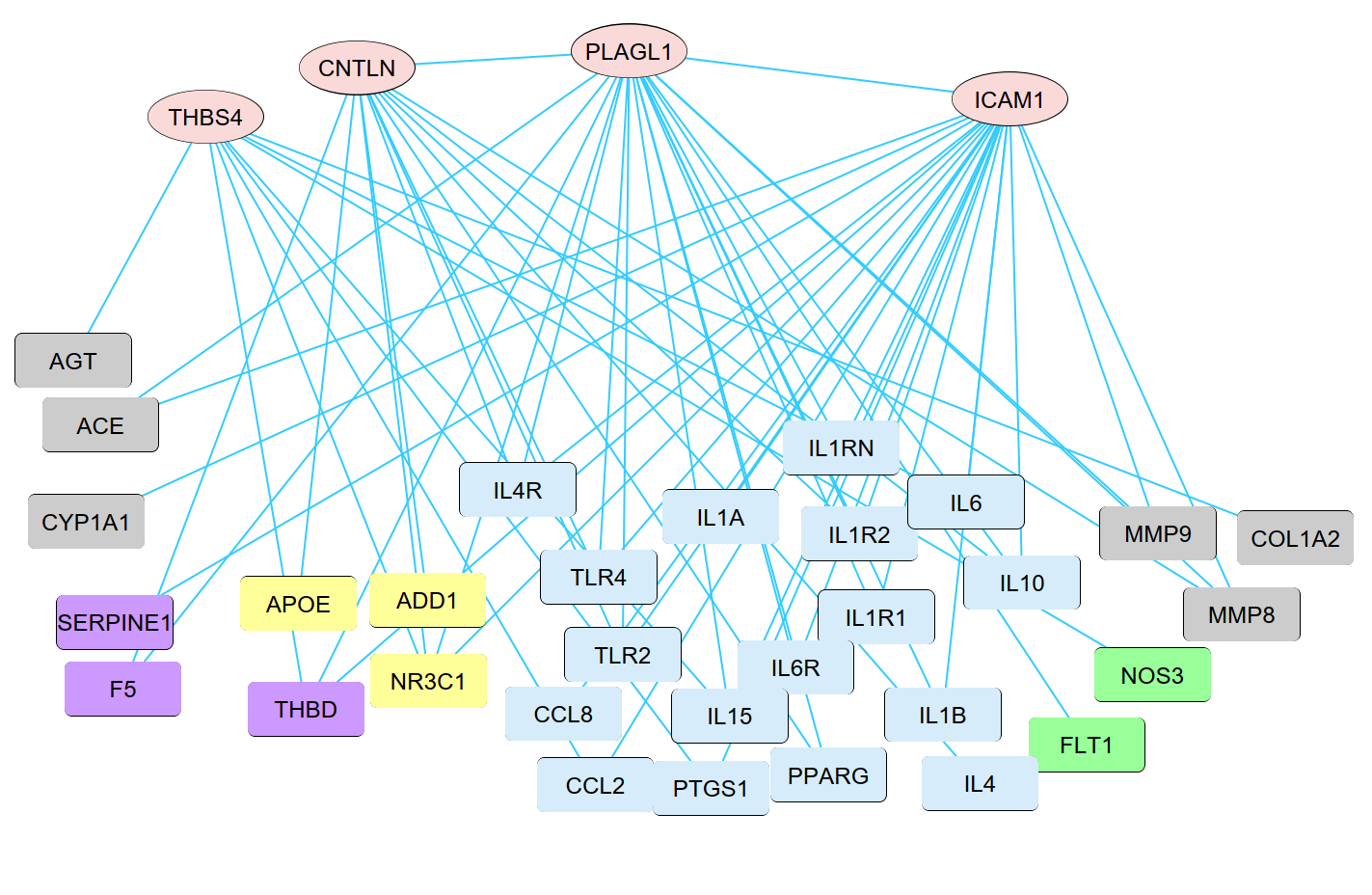

**Figure S11.** **Co-expression networks of the PTB-related genes in maternal and infant subtypes.** The four networks represent maternal PTB (a), infant PTB (b), BPD (c) and sepsis (d) of preterm infant, respectively. Oval nodes refer to the PTB related genes identified from the GWAS of the 1,744 embryo samples. Rectangle nodes refer to the 50-PTB markers derived from published datasets (see Addition file 1: Table S5) and involved in biological processes or functions, blue (immune or inflammation), yellow (apoptosis), green (angiogenesis), purple (coagulation) and grey (other). Co-expressive links in red (for maternal) or blue (for infant) between two nodes were based on Pearson correlative relations at p value < 0.01 significant level.
